## Supplementary material for "Predicting the impact of COVID-19 vaccination campaigns – a flexible age-dependent, spatially-stratified predictive model, accounting for multiple viral variants and vaccines": S1 Mathematical Appendix

### Appendix - Mathematical description

Here we present a concise formulation of the mathematical model, which is an extended SEIR model based on that underlying pandemic-preparedness-tool CovidSim 2.0 (cf. [1]) as well as the COVID-19 vaccination model in [2] and the COVID-19 risk group model in [3]. The presentation follows these references.

To accommodate age and spatial structure, different viral strains, and vaccination with different vaccine products, the population has to be partitioned into several compartments in the SEIR model. The model's flow chart is presented in S1 Figure. Figs 1 - 2 in the main text are simplified illustrations that facilitate understanding of the flow chart.

All model compartments are stratified by age and location, where we consider $s$ disjoint age classes (strata), indexed by $a$ ($a = 1, \ldots, s$), and $r$ different locations (regions), indexed by $l$ ($l = 1, \ldots, r$). We use $a$ and $l$ as super- or sub-scripts to indicate the age group and location, respectively. Furthermore, we assume that different variants of SARS-CoV-2 are transmitted in the population. The set of the SARS-CoV-2 variants is denoted by $M$. In the following we denote viral variants by $m \in M$, and use this notation in super- or sub-scripts. The set of available vaccine products is denoted by $V$. Particular vaccines are indexed by $v \in V$.

The model is also subdivided into several compartments that indicate the vaccination status of individuals. If we are not referring to a particular vaccination status, we use (.) as a placeholder in the super-scripts.

#### Model compartments

The model assumes that susceptible ($S$) individuals can be infected by one of the admissible SARS-CoV-2 variants (specified by the set $M$) by contacts with infected individuals. Co-infections with several SARS-CoV-2 variants are ignored. Individuals that recovered from an infection with any of the variants are assumed to be permanently immune against infection with the other variants. Susceptible individuals that become infected enter the latent phase ($E$), during which the infection is asymptomatic and the virus cannot yet be transmitted. Following the latent phase, individuals progress into the prodromal phase ($P$), during which they become infectious but remain asymptomatic. The infection progresses further into the fully-infectious phase ($I$), during which individuals are most infectious and symptoms might manifest. In the final late-infectious phase ($L$) individuals are less infectious than in the fully-infectious phase. From there, asymptomatic individuals will recover (and acquire permanent immunity against all SARS-CoV-2 variants), whereas symptomatic infections lead to either recovery or death.

A proportion of the population is unvaccinable (anti-vaxxers, individuals with contraindications, etc.). The remaining fraction of the population will get vaccinated – either as long as they are still susceptible or when they are already infected (we can ignore to model recovered individuals being vaccinated). The vaccine changes susceptibility to certain SARS-CoV-2 variants – reflecting that not every vaccine might protect against every viral variant.

To accommodate this, several model compartments have to be considered. In the following, we introduce these compartments and then describe the progression of the disease.

#### Susceptible individuals

Susceptible individuals (see Fig 2 in the main text, S1 Figure) are sub-divided by age group ($a$) and location ($l$) as well as into those that are: (i) waiting to be vaccinated ($S_{a,l}^{(\mathrm{U})}$); (ii) already vaccinated with vaccine $v$, for whom the outcome of the vaccination is still pending ($S_{a,l}^{(\mathrm{V},v)}$); (iii) already vaccinated with $v$ but only partially immunized ($S_{a,l}^{(\mathrm{PI},v)}$); (iv) not vaccinable (anti-vaxxers or individuals with contraindications) or vaccinated with $v$, but the vaccine completely failed to immunize ($S_{a,l}^{(\mathrm{NI})}$); or (v) that were immunized successfully by a vaccine, but might still be susceptible to certain variants ($R_{a,l}^{(\mathrm{Im},v)}$). Note, a vaccine $v$ might protect against all viral variants, in which case individuals within $R_{a,l}^{(\mathrm{Im},v)}$ are no longer susceptible but 'recovered'.

The waiting time for susceptibles ($S_{a,l}^{(\mathrm{U})}$) to get vaccinated is $D_{a,l}^{(\mathrm{V},v)}$, which is in general age and location dependent, reflecting the prioritization of vaccine distribution. Moreover, this parameter is time dependent, i.e., $D_{a,l}^{(\mathrm{V},v)} = D_{a,l}^{(\mathrm{V},v)}(t)$. Namely, vaccination does not start at the onset of the pandemic and the waiting time to get vaccinated depends on the availability of the vaccine products. This time dependence is omitted, except when necessary, to simplify the notation.

The effect of the vaccine is not immediate (pending immunization). Thus, individuals from $S_{a,l}^{(\mathrm{U})}$, if not infected, are moved into compartment ($S_{a,l}^{(\mathrm{V},v)}$) at rate $\nu_{a,l}^{(v)} = 1/D_{a,l}^{(\mathrm{V},v)}$. If not infected, after an average duration $D_a^{(v)}$, the outcome of the vaccine manifests: susceptibles either become (i) fully immunzed ($R_{a,l}^{(v)}$) with probability $f_S^{(\mathrm{Im},v)}$; (ii) partially immunized ($S_{a,l}^{(\mathrm{PI},v)}$, i.e., they can still get infected) with probability $f_S^{(\mathrm{PI},v)}$; or (iii) not immunized ($S_{a,l}^{(\mathrm{NI})}$, i.e., the vaccine had no effect) with probability $f_S^{(\mathrm{NI},v)}$. Hence, individuals (if not infected) are moved from $S_{a,l}^{(\mathrm{V},v)}$ to either $R_{a,l}^{(v)}$, $S_{a,l}^{(\mathrm{PI},v)}$, or $S_{a,l}^{(\mathrm{NI})}$ at rates $\alpha_a^{(v)} f_S^{(\mathrm{Im},v)}$, $\alpha_a^{(v)} f_S^{(\mathrm{PI},v)}$, or $\alpha_a^{(v)} f_S^{(\mathrm{NI},v)}$, respectively, where $\alpha_a^{(v)} = 1/D_a^{(v)}$. (Clearly, $f_S^{(\mathrm{Im},v)} + f_S^{(\mathrm{PI},v)} + f_S^{(\mathrm{NI},v)} = 1$.)

Individuals are assumed to be equally susceptible if they are waiting to be vaccinated ($S_{a,l}^{(\mathrm{U})}$), vaccinated but have pending immunity ($S_{a,l}^{(\mathrm{V},v)}$), or failed to immunize ($S_{a,l}^{(\mathrm{NI})}$). Individuals who have immunity against some but not all of the virus strains ($R_{a,l}^{(\mathrm{Im},v)}$) are still susceptible to some of the variants. Susceptibles that developed partial immunity ($S_{a,l}^{(\mathrm{PI},v)}$) are less likely to become infected than the other susceptibles. Namely, only fractions $p_P^{(m,v)}$, $p_I^{(m,v)}$, $p_L^{(m,v)}$ of contacts that would infect other susceptibles in the prodromal, fully-infectious, and late-infectious phases are infective, respectively. If infected, partial immunity manifests in a lower likelihood to develop symptoms (severe disease) and in lower mortality.

#### Infected individuals

The stages of the infection (latent, prodromal, fully infectious, and late infectious) are modeled by equivalent sub-states (Erlang states), through which individuals progress. Modeling these Erlang states yields more realistic model dynamics. Namely, the average duration spent in the various phases will no longer be exponentially but Erlang distributed (cf. [1, 3]). (Standard SEIR models implicitly assume exponential waiting times, hence the variance of the time spent in each phase of the disease is too high: the disease progresses too fast in some individuals and much slowly in others.)

If infected with variant $m$, unvaccinated individuals ($S_{a,l}^{(\mathrm{U})}$) first progress successively through the latent sub-states ($E_{k,a,l}^{(\mathrm{U},m)}$, $k = 1, \ldots, n_E^{(a,m)}$), where $n_E^{(a,m)}$ is the number of latent Erlang states. All unvaccinated individuals in age group $a$ and location $l$ infected with variant $m$ in the latent phase at time $t$ are

$$E_{\mathrm{Sum},a,l}^{(\mathrm{U},m)}(t) := \sum_{k=1}^{n_E^{(a,m)}} E_{k,a,l}^{(\mathrm{U},m)}(t). \tag{1a}$$

Similarly, the latent stages for individuals that are (i) vaccinated with $v$, for whom the outcome is pending, (ii) partially-immune, and (iii) unimmunized (including unvaccinable) individuals are denoted by $E_{k,a,l}^{(\mathrm{V},m,v)}$, $E_{k,a,l}^{(\mathrm{PI},m,v)}$, $E_{k,a,l}^{(\mathrm{NI},m)}$ ($k = 1, \ldots, n_E^{(a,m)}$), respectively. The total numbers of these individuals at time $t$ are, respectively,

$$E_{\mathrm{Sum},a,l}^{(\mathrm{V},m,v)}(t) := \sum_{k=1}^{n_E^{(a,m)}} E_{k,a,l}^{(\mathrm{V},m,v)}(t), \tag{1b}$$

$$E_{\mathrm{Sum},a,l}^{(\mathrm{PI},m,v)}(t) := \sum_{k=1}^{n_E^{(a,m)}} E_{k,a,l}^{(\mathrm{PI},m,v)}(t), \tag{1c}$$

and

$$E_{\mathrm{Sum},a,l}^{(\mathrm{NI},m)}(t) := \sum_{k=1}^{n_E^{(a,m)}} E_{k,a,l}^{(\mathrm{NI},m)}(t). \tag{1d}$$

Note that all unimmunized individuals, in age group $a$ and location $l$ that were infected with variant $m$ are modeled by the same compartments $E_{k,a,l}^{(\mathrm{NI},m)}$, i.e., it is irrelevant to specify which vaccine $v$ failed to immunize.

Following the latent phase, infected individuals progress into the prodromal phase. The prodromal phase is modeled similarly as the latent phase. The Erlang stages for prodromal individuals which are unvaccinated, vaccinated with pending vaccine outcome, partially-immune and not-immunized individuals are, respectively, denoted by $P_{k,a,l}^{(\mathrm{U},m)}$, $P_{k,a,l}^{(\mathrm{V},m,v)}$, $P_{k,a,l}^{(\mathrm{PI},m,v)}$, and $P_{k,a,l}^{(\mathrm{NI},m)}$ for $k = 1, \ldots n_P^{(a,m)}$. The total number of prodromal individuals in the different categories at time $t$ are

$$P^{(\mathrm{U},m)}_{\mathrm{Sum},a,l}(t) := \sum_{k=1}^{n_P^{(a,m)}} P^{(\mathrm{U},m)}_{k,a,l}(t), \tag{1e}$$

$$P^{(\mathrm{V},m,v)}_{\mathrm{Sum},a,l}(t) := \sum_{k=1}^{n_P^{(a,m)}} P^{(\mathrm{V},m,v)}_{k,a,l}(t), \tag{1f}$$

$$P^{(\mathrm{PI},m,v)}_{\mathrm{Sum},a,l}(t) := \sum_{k=1}^{n_P^{(a,m)}} P^{(\mathrm{PI},m,v)}_{k,a,l}(t), \tag{1g}$$

$$P^{(\mathrm{NI},m)}_{\mathrm{Sum},a,l}(t) := \sum_{k=1}^{n_P^{(a,m)}} P^{(\mathrm{NI},m)}_{k,a,l}(t). \tag{1h}$$

Similarly, the respective sub-states in the fully-infectious states are denoted by $I^{(\mathrm{U},m)}_{k,a,l}$, $I^{(\mathrm{V},m,v)}_{k,a,l}$, $I^{(\mathrm{PI},m,v)}_{k,a,l}$, $I^{(\mathrm{NI},m)}_{k,a,l}$ for $k = 1, \dots n_I^{(a,m)}$, and by $L^{(\mathrm{U},m)}_{k,a,l}$, $L^{(\mathrm{V},m,v)}_{k,a,l}$, $L^{(\mathrm{PI},m,v)}_{k,a,l}$, $L^{(\mathrm{NI},m)}_{k,a,l}$ ($k = 1, \dots, n_L^{(a,m)}$) for the late-infectious stages. The total numbers of fully-infectious individuals in the respective categories are

$$I^{(\mathrm{U},m)}_{\mathrm{Sum},a,l}(t) := \sum_{k=1}^{n_I^{(a,m)}} I^{(\mathrm{U},m)}_{k,a,l}(t), \tag{1i}$$

$$I^{(\mathrm{V},m,v)}_{\mathrm{Sum},a,l}(t) := \sum_{k=1}^{n_I^{(a,m)}} I^{(\mathrm{V},m,v)}_{k,a,l}(t), \tag{1j}$$

$$I^{(\mathrm{PI},m,v)}_{\mathrm{Sum},a,l}(t) := \sum_{k=1}^{n_I^{(a,m)}} I^{(\mathrm{PI},m,v)}_{k,a,l}(t), \tag{1k}$$

$$I^{(\mathrm{NI},m)}_{\mathrm{Sum},a,l}(t) := \sum_{k=1}^{n_I^{(a,m)}} I^{(\mathrm{NI},m)}_{k,a,l}(t), \tag{1l}$$

and those of the late-infectious individuals are

$$L^{(\mathrm{U},m)}_{\mathrm{Sum},a,l}(t) := \sum_{k=1}^{n_L^{(a,m)}} L^{(\mathrm{U},m)}_{k,a,l}(t), \tag{1m}$$

$$L^{(\mathrm{V},m,v)}_{\mathrm{Sum},a,l}(t) := \sum_{k=1}^{n_L^{(a,m)}} L^{(\mathrm{V},m,v)}_{k,a,l}(t), \tag{1n}$$

$$L^{(\mathrm{PI},m,v)}_{\mathrm{Sum},a,l}(t) := \sum_{k=1}^{n_L^{(a,m)}} L^{(\mathrm{PI},m,v)}_{k,a,l}(t), \tag{1o}$$

$$L^{(\mathrm{NI},m)}_{\mathrm{Sum},a,l}(t) := \sum_{k=1}^{n_L^{(a,m)}} L^{(\mathrm{NI},m)}_{k,a,l}(t). \tag{1p}$$

#### Course of the infection, recovery or death

Once infected with variant $m$, the average duration of the latent, prodromal, fully-infectious, and late-infectious periods for individuals in age-group $a$ are $D_E^{(a,m)}$,

$D_P^{(a,m)}$, $D_I^{(a,m)}$, and $D_L^{(a,m)}$, respectively. In each of these phases individuals transit through the equivalent sub-states (Erlang states). Individuals in age group $a$ infected with variant $m$ leave each latent sub-state at rate

$$\varepsilon_{a,m} := \frac{n_E^{(a,m)}}{D_E^{(a,m)}}. \tag{2a}$$

Similarly, the prodromal, fully-infectious, and late-infectious sub-states are left, respectively, at rates

$$\varphi_{a,m} := \frac{n_P^{(a,m)}}{D_P^{(a,m)}}, \gamma_{a,m} := \frac{n_I^{(a,m)}}{D_I^{(a,m)}}, \text{ and } \delta_{a,m} := \frac{n_L^{(a,m)}}{D_L^{(a,m)}}. \tag{2b}$$

Individuals are not yet infectious or show symptoms in the latent phase (see Fig 1). They still show no symptoms in the prodromal phase but are already infectious – however, not to the full extent. At the beginning of the fully-infectious period individuals either stay asymptomatic or become symptomatic, i.e., they become sick. A fraction $f_{\text{Sick}}^{(a,m)}$ of unvaccinated individuals, vaccinated individuals with pending vaccination outcome, and unimmunized individuals gets sick. Partially-immune individuals are less likely to develop symptoms, namely a fraction $f_{\text{Sick}}^{(\text{PI},a,m,v)} \leq f_{\text{Sick}}^{(a,m)}$ develops symptoms.

The number of symptomatic individuals per age group and location that are infected with variant $m$ is

$$I_{\text{Sick},a,l}^{(\text{U},m)}(t) := f_{\text{Sick}}^{(a,m)} I_{\text{Sum},a,l}^{(\text{U},m)}(t), \tag{3a}$$

among individuals waiting to be vaccinated,

$$I_{\text{Sick},a,l}^{(\text{V},m)}(t) := f_{\text{Sick}}^{(a,m)} \sum_{v \in V} I_{\text{Sum},a,l}^{(\text{V},m,v)}(t), \tag{3b}$$

among individuals with pending effect of the vaccination,

$$I_{\text{Sick},a,l}^{(\text{PI},m)}(t) := \sum_{v \in V} f_{\text{Sick}}^{(\text{PI},a,m,v)} I_{\text{Sum},a,l}^{(\text{PI},m,v)}(t), \tag{3c}$$

among individuals that are partially-immune, and

$$I_{\text{Sick},a,l}^{(\text{NI},m)}(t) := f_{\text{Sick}}^{(a,m)} I_{\text{Sum},a,l}^{(\text{NI},m)}(t), \tag{3d}$$

for unvaccinable individuals and those that failed to immunize. For partially-immune individuals it is important to also stratify the number of sick individuals by vaccine. We define

$$I_{\text{Sick},a,l}^{(\text{PI},m,v)}(t) := f_{\text{Sick}}^{(\text{PI},a,m,v)} I_{\text{Sum},a,l}^{(\text{PI},m,v)}(t). \tag{4}$$

Similarly, in the late-infectious phase the corresponding numbers are

$$L_{\text{Sick},a,l}^{(\text{U},m)}(t) := f_{\text{Sick}}^{(a,m)} L_{\text{Sum},a,l}^{(\text{U},m)}(t), \tag{5a}$$

$$L_{\text{Sick},a,l}^{(\text{V},m)}(t) := f_{\text{Sick}}^{(a,m)} \sum_{v \in V} L_{\text{Sum},a,l}^{(\text{V},m,v)}(t), \tag{5b}$$

$$L_{\text{Sick},a,l}^{(\text{PI},m)}(t) := \sum_{v \in V} f_{\text{Sick}}^{(\text{PI},a,m,v)} L_{\text{Sum},a,l}^{(\text{PI},m,v)}(t), \tag{5c}$$

and

$$L^{(\mathrm{NI},m)}_{\mathrm{Sick},a,l}(t) := f^{(a,m)}_{\mathrm{Sick}} L^{(\mathrm{NI},m)}_{\mathrm{Sum},a,l}(t). \tag{5d}$$

among individuals that failed to immunize. Also the symptomatic infections of partially-immune late-infectious individuals need to be stratified by vaccine, i.e.,

$$L^{(\mathrm{PI},m,v)}_{\mathrm{Sick},a,l}(t) := f^{(\mathrm{PI},a,m,v)}_{\mathrm{Sick}} L^{(\mathrm{PI},m,v)}_{\mathrm{Sum},a,l}(t). \tag{6}$$

Individuals ultimately recover from asymptomatic infections and progress from the last late-infectious sub-state to the respective compartment of recovered individuals. The number of recovered individuals in age group $a$ at location $l$ that recovered from infection with variant $m$ is denoted by $R^{(\mathrm{Inf},m)}_{a,l}$. A fraction $f^{(a,m)}_{\mathrm{Dead}}$ of symptomatic infections is lethal. This fraction is lower for partially-immune individuals $f^{(\mathrm{PI},a,m,v)}_{\mathrm{Dead}} \leq f^{(a,m)}_{\mathrm{Dead}}$.

##### Vaccination during infection

Susceptible individuals waiting to get vaccinated might actually be vaccinated when already infected. We assume that the vaccine will not change the course of the infection after the fully-infectious phase was already reached. Therefore, vaccination does not have to be explicitly modeled in those phases. If individuals are vaccinated during the latent or prodromal phase, they change their status and are moved into the corresponding compartments of latent and prodromal individuals with pending vaccination outcome. More precisely, individuals in compartments $E^{(\mathrm{U},m)}_{k,a,l}$ and $P^{(\mathrm{U},m)}_{k,a,l}$ are moved into compartments $E^{(\mathrm{V},m,v)}_{k,a,l}$ and $P^{(\mathrm{V},m,v)}_{k,a,l}$ at rate $\nu^{(v)}_{a,l}$.

The output of the vaccinated individuals might manifest in any of the latent, prodromal, fully-infectious, or late-infectious sub-states. The outcome of the vaccine can result in faster recovery $R^{(\mathrm{Inf},m)}_{a,l}$, in partial immunity protecting from severe infection and mortality, or in a failure to immunize. Vaccination results in early recovery in fractions $f^{(\mathrm{Im},v)}_{E}$, $f^{(\mathrm{Im},v)}_{P}$, $f^{(\mathrm{Im},v)}_{I}$, and $f^{(\mathrm{Im},v)}_{L}$ of the latent, prodromal, fully-, and late-infectious individuals, while fractions $f^{(\mathrm{PI},v)}_{E}$, $f^{(\mathrm{PI},v)}_{P}$, $f^{(\mathrm{PI},v)}_{I}$, and $f^{(\mathrm{PI},v)}_{L}$ result in partial immunity. The vaccine results in no immunization for the remaining fractions ($f^{(\mathrm{NI},v)}_{E}$, $f^{(\mathrm{NI},v)}_{P}$, $f^{(\mathrm{NI},v)}_{I}$, and $f^{(\mathrm{NI},v)}_{L}$). (Clearly, $f^{(\mathrm{Im},v)}_{E} + f^{(\mathrm{PI},v)}_{E} + f^{(\mathrm{NI},v)}_{E} = 1$, $f^{(\mathrm{Im},v)}_{P} + f^{(\mathrm{PI},v)}_{P} + f^{(\mathrm{NI},v)}_{P} = 1$, $f^{(\mathrm{Im},v)}_{I} + f^{(\mathrm{PI},v)}_{I} + f^{(\mathrm{NI},v)}_{I} = 1$, and $f^{(\mathrm{Im},v)}_{L} + f^{(\mathrm{PI},v)}_{L} + f^{(\mathrm{NI},v)}_{L} = 1$.)

#### Contact rate

The basic reproduction number varies between viral variants. Let $R^{(m)}_0$ be the basic reproduction number of variant $m$, which is assumed to fluctuate seasonally around its annual average, $\bar{R}^{(m)}_0$, by an amplitude $A$ ($0 \leq A \leq 1$) with a peak at time $t_{R_{0_{\max}}}$, i.e.,

$$R^{(m)}_0(t) := \bar{R}^{(m)}_0 \left(1 + A \cos\left(2\pi \frac{t - t_{R_{0_{\max}}}}{365}\right)\right). \tag{7}$$

Note that the amount of seasonal fluctuations and the time $R^{(m)}_0$ peaks, are independent of the viral variant $m$, as this reflects extrinsic factors such as climate.

Classically, the basic reproduction number is the average number of infections caused by a single infected individual in a completely susceptible population, in which no interventions occur (cf. [4]). Here, we are considering an age- and spatially-structured population, i.e., the population consists of heterogeneous sub-populations. In such a setting, $\bar{R}_0^{(m)}$ is determined by the contact behaviors via the next generation matrix (cf. see [4]). Importantly, encounters between individuals are not random. The likelihood that two individuals encounter depends on the age strata and locations of the two individuals. Let $x_{a',l'}^{(a,l)}(t)$ be the factor that mediates random encounters between individuals in age group $a$ in location $l$ with individuals in age group $a'$ in location $l'$ at time $t$. This yields a time-dependent symmetric mixing matrix $X(t)$. This matrix corrects for the true contact behavior and is described in detail below (see below section "Mathematical description of the demographic mixing matrix"). The mixing matrix is time dependent because the contact behavior is influenced by general contact reducing interventions, e.g., curfews etc., described in detail below.

The relative contagiousness of individuals infected with variant $m$ in age group $a$ in the prodromal, fully-infectious and late-infectious states are $c_P^{(m)}$, $c_I^{(m)}$, $c_L^{(m)}$. The rates of infective contacts with individuals infected with variant $m$ are determined by multiplying the mixing matrix with the following "contact rates":

$$\beta_P^{(m)}(t) := c_P^{(m)} \bar{R}_0^{(m,\text{adj})} \left(1 + A\cos\left(2\pi \frac{t - t_{R_{0\max}}}{365}\right)\right), \tag{8a}$$

$$\beta_I^{(m)}(t) := c_I^{(m)} \bar{R}_0^{(m,\text{adj})} \left(1 + A\cos\left(2\pi \frac{t - t_{R_{0\max}}}{365}\right)\right), \tag{8b}$$

$$\beta_L^{(m)}(t) := c_L^{(m)} \bar{R}_0^{(m,\text{adj})} \left(1 + A\cos\left(2\pi \frac{t - t_{R_{0\max}}}{365}\right)\right), \tag{8c}$$

where $\bar{R}_0^{(m,\text{adj})}$ is the adjusted average reproduction number that accounts for the contact behavior in the population.

The effective number of individuals that can infect susceptibles is determined by case isolation (i.e., quarantine or home isolation). The contact rates are mediated additionally by general contact reduction.

#### Case isolation

During the time interval $[t_{\text{Iso}_1}, t_{\text{Iso}_2}]$, a fraction $f_{\text{Iso}}^{(l)}$ of individuals with symptomatic infections that seek medical help will be isolated in quarantine wards until the wards are full, in which case they are sent into home isolation. The fraction of individuals being isolated and the maximum capacity, $Q_{\max}^{(l)}$, of the quarantine wards are location dependent. Quarantine wards guarantee perfect isolation, whereas home isolation reduces only a fraction $p_{\text{Home}}$ of contacts. The total number of individuals in location $l$ that is isolated in quarantine wards or at home at time $t$ is

$$\begin{aligned} Q_l(t) := f_{\text{Iso}}^{(l)} \sum_{a=1}^{s} \sum_{m\in M} \Big( & I_{\text{Sick},a,l}^{(\text{U},m)}(t) + L_{\text{Sick},a,l}^{(\text{U},m)}(t) + I_{\text{Sick},a,l}^{(\text{V},m)}(t) + L_{\text{Sick},a,l}^{(\text{V},m)}(t) \\ & + I_{\text{Sick},a,l}^{(\text{PI},m)}(t) + L_{\text{Sick},a,l}^{(\text{PI},m)}(t) + I_{\text{Sick},a,l}^{(\text{NI},m)}(t) + L_{\text{Sick},a,l}^{(\text{NI},m)}(t) \Big). \end{aligned} \tag{9}$$

The total numbers of quarantined fully-infectious individuals, waiting to be vaccinated, whose immunization is pending (V), that are unvaccinable or failed to immunized (NI), or that were partially immunized (PI) are

$$I^{(\mathrm{U},m)}_{\mathrm{Iso},a,l} := \begin{cases} f^{(l)}_{\mathrm{Iso}} I^{(\mathrm{U},m)}_{\mathrm{Sick},a,l} & \text{if } t_{\mathrm{Iso}_1} \le t \le t_{\mathrm{Iso}_2} \text{ and } Q_l(t) \le Q^{(l)}_{\max}, \\ f^{(l)}_{\mathrm{Iso}} I^{(\mathrm{U},m)}_{\mathrm{Sick},a,l} \dfrac{Q^{(l)}_{\max}}{Q_l(t)} & \text{if } t_{\mathrm{Iso}_1} \le t \le t_{\mathrm{Iso}_2} \text{ and } Q_l(t) > Q^{(l)}_{\max}, \\ 0 & \text{otherwise,} \end{cases} \tag{10a}$$

$$I^{(\mathrm{V},m)}_{\mathrm{Iso},a,l} := \begin{cases} f^{(l)}_{\mathrm{Iso}} I^{(\mathrm{V},m)}_{\mathrm{Sick},a,l} & \text{if } t_{\mathrm{Iso}_1} \le t \le t_{\mathrm{Iso}_2} \text{ and } Q_l(t) \le Q^{(l)}_{\max}, \\ f^{(l)}_{\mathrm{Iso}} I^{(\mathrm{V},m)}_{\mathrm{Sick},a,l} \dfrac{Q^{(l)}_{\max}}{Q_l(t)} & \text{if } t_{\mathrm{Iso}_1} \le t \le t_{\mathrm{Iso}_2} \text{ and } Q_l(t) > Q^{(l)}_{\max}, \\ 0 & \text{otherwise,} \end{cases} \tag{10b}$$

$$I^{(\mathrm{PI},m,v)}_{\mathrm{Iso},a,l} := \begin{cases} f^{(l)}_{\mathrm{Iso}} I^{(\mathrm{PI},m,v)}_{\mathrm{Sick},a,l} & \text{if } t_{\mathrm{Iso}_1} \le t \le t_{\mathrm{Iso}_2} \text{ and } Q_l(t) \le Q^{(l)}_{\max}, \\ f^{(l)}_{\mathrm{Iso}} I^{(\mathrm{PI},m,v)}_{\mathrm{Sick},a,l} \dfrac{Q^{(l)}_{\max}}{Q_l(t)} & \text{if } t_{\mathrm{Iso}_1} \le t \le t_{\mathrm{Iso}_2} \text{ and } Q_l(t) > Q^{(l)}_{\max}, \\ 0 & \text{otherwise,} \end{cases} \tag{10c}$$

and

$$I^{(\mathrm{NI},m)}_{\mathrm{Iso},a,l} := \begin{cases} f^{(l)}_{\mathrm{Iso}} I^{(\mathrm{NI},m)}_{\mathrm{Sick},a,l} & \text{if } t_{\mathrm{Iso}_1} \le t \le t_{\mathrm{Iso}_2} \text{ and } Q_l(t) \le Q^{(l)}_{\max}, \\ f^{(l)}_{\mathrm{Iso}} I^{(\mathrm{NI},m)}_{\mathrm{Sick},a,l} \dfrac{Q^{(l)}_{\max}}{Q_l(t)} & \text{if } t_{\mathrm{Iso}_1} \le t \le t_{\mathrm{Iso}_2} \text{ and } Q_l(t) > Q^{(l)}_{\max}, \\ 0 & \text{otherwise.} \end{cases} \tag{10d}$$

Note, the number of individuals in isolation is stratified by vaccine only for partially-immune individuals. Furthermore, the respective numbers of fully-infectious individuals in home isolation are

$$I^{(\mathrm{U},m)}_{\mathrm{Home},a,l} := \begin{cases} I^{(\mathrm{U},m)}_{\mathrm{Sick},a,l} f^{(l)}_{\mathrm{Iso}} \left(1 - \dfrac{Q^{(l)}_{\max}}{Q_l(t)}\right) & \text{if } t_{\mathrm{Iso}_1} \le t \le t_{\mathrm{Iso}_2} \text{ and } Q_l(t) > Q^{(l)}_{\max}, \\ 0 & \text{otherwise,} \end{cases} \tag{11a}$$

$$I^{(\mathrm{V},m)}_{\mathrm{Home},a,l} := \begin{cases} I^{(\mathrm{V},m)}_{\mathrm{Sick},a,l} f^{(l)}_{\mathrm{Iso}} \left(1 - \dfrac{Q^{(l)}_{\max}}{Q_l(t)}\right) & \text{if } t_{\mathrm{Iso}_1} \le t \le t_{\mathrm{Iso}_2} \text{ and } Q_l(t) > Q^{(l)}_{\max}, \\ 0 & \text{otherwise,} \end{cases} \tag{11b}$$

$$I^{(\mathrm{PI},m,v)}_{\mathrm{Home},a,l} := \begin{cases} I^{(\mathrm{PI},m,v)}_{\mathrm{Sick},a,l} f^{(l)}_{\mathrm{Iso}} \left(1 - \dfrac{Q^{(l)}_{\max}}{Q_l(t)}\right) & \text{if } t_{\mathrm{Iso}_1} \le t \le t_{\mathrm{Iso}_2} \text{ and } Q_l(t) > Q^{(l)}_{\max}, \\ 0 & \text{otherwise,} \end{cases} \tag{11c}$$

and

$$I^{(\mathrm{NI},m)}_{\mathrm{Home},a,l} := \begin{cases} I^{(\mathrm{NI},m)}_{\mathrm{Sick},a,l} f^{(l)}_{\mathrm{Iso}} \left(1 - \dfrac{Q^{(l)}_{\max}}{Q_l(t)}\right) & \text{if } t_{\mathrm{Iso}_1} \le t \le t_{\mathrm{Iso}_2} \text{ and } Q_l(t) > Q^{(l)}_{\max}, \\ 0 & \text{otherwise.} \end{cases} \tag{11d}$$

The effective numbers of fully-infectious individuals in the various groups subject to isolation that can infect susceptibles become

$$I^{(\mathrm{U},m)}_{\mathrm{Eff},a,l} := I^{(\mathrm{U},m)}_{\mathrm{Sum},a,l} - I^{(\mathrm{U},m)}_{\mathrm{Iso},a,l} - p_{\mathrm{Home}} I^{(\mathrm{U},m)}_{\mathrm{Home},a,l}\,, \tag{12a}$$

$$I^{(\mathrm{V},m)}_{\mathrm{Eff},a,l} := \sum_{v\in V} I^{(\mathrm{V},m,v)}_{\mathrm{Sum},a,l} - I^{(\mathrm{V},m)}_{\mathrm{Iso},a,l} - p_{\mathrm{Home}} I^{(\mathrm{V},m)}_{\mathrm{Home},a,l}\,, \tag{12b}$$

$$I^{(\mathrm{PI},m,v)}_{\mathrm{Eff},a,l} := I^{(\mathrm{PI},m,v)}_{\mathrm{Sum},a,l} - I^{(\mathrm{PI},m,v)}_{\mathrm{Iso},a,l} - p_{\mathrm{Home}} I^{(\mathrm{PI},m,v)}_{\mathrm{Home},a,l}\,, \tag{12c}$$

$$I^{(\mathrm{NI},m)}_{\mathrm{Eff},a,l} := I^{(\mathrm{NI},m)}_{\mathrm{Sum},a,l} - I^{(\mathrm{NI},m)}_{\mathrm{Iso},a,l} - p_{\mathrm{Home}} I^{(\mathrm{NI},m)}_{\mathrm{Home},a,l}. \tag{12d}$$

The corresponding numbers of late-infectious individuals in quarantine are

$$L^{(\mathrm{U},m)}_{\mathrm{Iso},a,l} := \begin{cases} f^{(l)}_{\mathrm{Iso}} L^{(\mathrm{U},m)}_{\mathrm{Sick},a,l} & \text{if } t_{\mathrm{Iso}_1} \le t \le t_{\mathrm{Iso}_2} \text{ and } Q_l(t) \le Q^{(l)}_{\max}, \\ f^{(l)}_{\mathrm{Iso}} L^{(\mathrm{U},m)}_{\mathrm{Sick},a,l} \dfrac{Q^{(l)}_{\max}}{Q_l(t)} & \text{if } t_{\mathrm{Iso}_1} \le t \le t_{\mathrm{Iso}_2} \text{ and } Q_l(t) > Q^{(l)}_{\max}, \\ 0 & \text{otherwise,} \end{cases} \tag{13a}$$

$$L^{(\mathrm{V},m)}_{\mathrm{Iso},a,l} := \begin{cases} f^{(l)}_{\mathrm{Iso}} L^{(\mathrm{V},m)}_{\mathrm{Sick},a,l} & \text{if } t_{\mathrm{Iso}_1} \le t \le t_{\mathrm{Iso}_2} \text{ and } Q_l(t) \le Q^{(l)}_{\max}, \\ f^{(l)}_{\mathrm{Iso}} L^{(\mathrm{V},m)}_{\mathrm{Sick},a,l} \dfrac{Q^{(l)}_{\max}}{Q_l(t)} & \text{if } t_{\mathrm{Iso}_1} \le t \le t_{\mathrm{Iso}_2} \text{ and } Q_l(t) > Q^{(l)}_{\max}, \\ 0 & \text{otherwise,} \end{cases} \tag{13b}$$

$$L^{(\mathrm{PI},m,v)}_{\mathrm{Iso},a,l} := \begin{cases} f^{(l)}_{\mathrm{Iso}} L^{(\mathrm{PI},m,v)}_{\mathrm{Sick},a,l} & \text{if } t_{\mathrm{Iso}_1} \le t \le t_{\mathrm{Iso}_2} \text{ and } Q_l(t) \le Q^{(l)}_{\max}, \\ f^{(l)}_{\mathrm{Iso}} L^{(\mathrm{PI},m,v)}_{\mathrm{Sick},a,l} \dfrac{Q^{(l)}_{\max}}{Q_l(t)} & \text{if } t_{\mathrm{Iso}_1} \le t \le t_{\mathrm{Iso}_2} \text{ and } Q_l(t) > Q^{(l)}_{\max}, \\ 0 & \text{otherwise,} \end{cases} \tag{13c}$$

and

$$L^{(\mathrm{NI},m)}_{\mathrm{Iso},a,l} := \begin{cases} f^{(l)}_{\mathrm{Iso}} L^{(\mathrm{NI},m)}_{\mathrm{Sick},a,l} & \text{if } t_{\mathrm{Iso}_1} \le t \le t_{\mathrm{Iso}_2} \text{ and } Q_l(t) \le Q^{(l)}_{\max}, \\ f^{(l)}_{\mathrm{Iso}} L^{(\mathrm{NI},m)}_{\mathrm{Sick},a,l} \dfrac{Q^{(l)}_{\max}}{Q_l(t)} & \text{if } t_{\mathrm{Iso}_1} \le t \le t_{\mathrm{Iso}_2} \text{ and } Q_l(t) > Q^{(l)}_{\max}, \\ 0 & \text{otherwise,} \end{cases} \tag{13d}$$

while those in home isolation are

$$L^{(\mathrm{U},m)}_{\mathrm{Home},a,l} := \begin{cases} L^{(\mathrm{U},m)}_{\mathrm{Sick},a,l} f^{(l)}_{\mathrm{Iso}} \Big(1 - \dfrac{Q^{(l)}_{\max}}{Q_l(t)}\Big) & \text{if } t_{\mathrm{Iso}_1} \le t \le t_{\mathrm{Iso}_2} \text{ and } Q_l(t) > Q^{(l)}_{\max}, \\ 0 & \text{otherwise,} \end{cases} \tag{14a}$$

$$L^{(\mathrm{V},m)}_{\mathrm{Home},a,l} := \begin{cases} L^{(\mathrm{V},m)}_{\mathrm{Sick},a,l} f^{(l)}_{\mathrm{Iso}} \Big(1 - \dfrac{Q^{(l)}_{\max}}{Q_l(t)}\Big) & \text{if } t_{\mathrm{Iso}_1} \le t \le t_{\mathrm{Iso}_2} \text{ and } Q_l(t) > Q^{(l)}_{\max}, \\ 0 & \text{otherwise,} \end{cases} \tag{14b}$$

$$L^{(\mathrm{PI},m,v)}_{\mathrm{Home},a,l} := \begin{cases} L^{(\mathrm{PI},m,v)}_{\mathrm{Sick},a,l} f^{(l)}_{\mathrm{Iso}} \Big(1 - \dfrac{Q^{(l)}_{\max}}{Q_l(t)}\Big) & \text{if } t_{\mathrm{Iso}_1} \le t \le t_{\mathrm{Iso}_2} \text{ and } Q_l(t) > Q^{(l)}_{\max}, \\ 0 & \text{otherwise,} \end{cases} \tag{14c}$$

and

$$L^{(\mathrm{NI},m)}_{\mathrm{Home},a,l} := \begin{cases} L^{(\mathrm{NI},m)}_{\mathrm{Sick},a,l} f^{(l)}_{\mathrm{Iso}} \Big(1 - \dfrac{Q^{(l)}_{\max}}{Q_l(t)}\Big) & \text{if } t_{\mathrm{Iso}_1} \le t \le t_{\mathrm{Iso}_2} \text{ and } Q_l(t) > Q^{(l)}_{\max}, \\ 0 & \text{otherwise.} \end{cases} \tag{14d}$$

The effective numbers of late-infectious individuals in those groups subject to isolation that can infect susceptibles becomes

$$L^{(\mathrm{U},m)}_{\mathrm{Eff},a,l} := L^{(\mathrm{U},m)}_{\mathrm{Sum},a,l} - L^{(\mathrm{U},m)}_{\mathrm{Iso},a,l} - p_{\mathrm{Home}} L^{(\mathrm{U},m)}_{\mathrm{Home},a,l}\,, \tag{15a}$$

$$L^{(\mathrm{V},m)}_{\mathrm{Eff},a,l} := \sum_{v \in V} L^{(\mathrm{V},m,v)}_{\mathrm{Sum},a,l} - L^{(\mathrm{V},m)}_{\mathrm{Iso},a,l} - p_{\mathrm{Home}} L^{(\mathrm{V},m)}_{\mathrm{Home},a,l}\,, \tag{15b}$$

$$L^{(\mathrm{PI},m,v)}_{\mathrm{Eff},a,l} := L^{(\mathrm{PI},m,v)}_{\mathrm{Sum},a,l} - L^{(\mathrm{PI},m,v)}_{\mathrm{Iso},a,l} - p_{\mathrm{Home}} L^{(\mathrm{PI},m,v)}_{\mathrm{Home},a,l}\,, \tag{15c}$$

$$L^{(\mathrm{NI},m)}_{\mathrm{Eff},a,l} := L^{(\mathrm{NI},m)}_{\mathrm{Sum},a,l} - L^{(\mathrm{NI},m)}_{\mathrm{Iso},a,l} - p_{\mathrm{Home}} L^{(\mathrm{NI},m)}_{\mathrm{Home},a,l}. \tag{15d}$$

#### Force of infection

The contagiousness of partially-immune infected individuals, vaccinated with vaccine $v$ and infected with variant $m$ reduces by a fraction $p^{(m,v)}_P$, $p^{(m,v)}_I$, $p^{(m,v)}_L$ during the prodromal, fully-infectious, and late-infectious periods. Taking the contact rates (8), general contact reduction reflected by the contact adjustments $x^{(a,l)}_{a',l'}(t)$, and the numbers of infectious individuals that effectively participate in infection (12), (15) into account, the (internal) force of infection by variant $m$ experienced by susceptible unprotected by a vaccine is

$$\begin{aligned} \lambda^{(m)}_{a,l}(t) := \sum_{a'=1}^{s} \sum_{l'=1}^{r} x^{(a,l)}_{a',l'}(t) \Bigg( & \beta^{(m)}_P(t) \Big( P^{(\mathrm{U},m)}_{\mathrm{Sum},a',l'}(t) + P^{(\mathrm{NI},m)}_{\mathrm{Sum},a',l'}(t) \\ & \quad + \sum_{v \in V} \Big( P^{(\mathrm{V},m,v)}_{\mathrm{Sum},a',l'}(t) + (1 - p^{(m,v)}_P) P^{(\mathrm{PI},m,v)}_{\mathrm{Sum},a',l'}(t) \Big) \Big) \\ & + \beta^{(m)}_I(t) \Big( I^{(\mathrm{U},m)}_{\mathrm{Eff},a,l}(t) + I^{(\mathrm{V},m)}_{\mathrm{Eff},a',l'}(t) + I^{(\mathrm{NI},m)}_{\mathrm{Eff},a',l'}(t) \\ & \quad + \sum_{v \in V} (1 - p^{(m,v)}_I) I^{(\mathrm{PI},m,v)}_{\mathrm{Eff},a',l'}(t) \Big) \\ & + \beta^{(m)}_L(t) \Big( L^{(\mathrm{U},m)}_{\mathrm{Eff},a',l'}(t) + L^{(\mathrm{V},m)}_{\mathrm{Eff},a',l'}(t) + L^{(\mathrm{NI},m)}_{\mathrm{Eff},a',l'}(t) \\ & \quad + \sum_{v \in V} (1 - p^{(m,v)}_L) L^{(\mathrm{PI},m,v)}_{\mathrm{Eff},a',l'}(t) \Big) \Bigg) \\ & + \lambda^{(a,l,m)}_{\mathrm{Ext}}(t), \end{aligned} \tag{16}$$

where $\lambda_{\text{Ext}}^{(a,l,m)}(t)$ is the external force of infection with variant $m$ at time $t$ experienced by individuals in location $l$ and age group $a$, i.e., resulting from contacts with infected individuals from outside the population.

The force of infection experienced by individuals that were partially immunized with vaccine $v$ is given by

$$\lambda_{a,l}^{(\text{PI},m,v)}(t) := g(m,v)\lambda_{a,l}^{(m)}(t), \tag{17}$$

where the function $g(m,v)$ determines the fraction by which the force of infection by variant $m$ is reduced if an individual is partially immunized with vaccine $v$. Individuals that were successfully immunized by vaccination against one or more variants might still be infected by some other variants. The force of infection experienced by these individuals is

$$\lambda_{a,l}^{(\text{Im},m,v)}(t) := h(m,v)\lambda_{a,l}^{(m)}(t), \tag{18}$$

where the function $h(m,v)$ determines the fraction by which the force of infection by variant $m$ is reduced if an individual is immunized with vaccine $v$. Clearly $h(m,v) = 0$ for all variants $m$ against which vaccine $v$ was successfully immunizing.

#### Dynamics of susceptibles

The change in the number of susceptibles in age class $a$ and location $l$ waiting to be vaccinated is determined by the forces of infection of the various viral variants and the rates at which these susceptibles are vaccinated with the different vaccine products. It is given by

$$\frac{dS_{a,l}^{(\text{U})}(t)}{dt} = -\frac{S_{a,l}^{(\text{U})}(t)}{N}\sum_{m\in M}\lambda_{a,l}^{(m)}(t) - S_{a,l}^{(\text{U})}(t)\sum_{v\in V}\nu_{a,l}^{(v)}. \tag{19a}$$

Changes in the number of susceptibles vaccinated with $v$ with pending vaccination outcome are determined by the rate of vaccination with $v$, the rate at which the vaccination outcome manifests, and the forces of infection, i.e.,

$$\frac{dS_{a,l}^{(\text{V},v)}(t)}{dt} = S_{a,l}^{(\text{U})}(t)\nu_{a,l}^{(v)} - \alpha_a^{(v)}S_{a,l}^{(\text{V},v)}(t) - \frac{S_{a,l}^{(\text{V},v)}(t)}{N}\sum_{m\in M}\lambda_{a,l}^{(m)}(t). \tag{19b}$$

A fraction of susceptibles vaccinated with $v$ is developing partial immunity at rate $\alpha_a^{(v)}$. These individuals can still be infected, however a reduced force of infection (depending on variant $m$ and vaccine $v$) is acting on them. The number of partially-immune susceptibles changes hence according to

$$\begin{aligned}\frac{dS_{a,l}^{(\text{PI},v)}(t)}{dt} &= \alpha_a^{(v)} f_S^{(\text{PI},v)} S_{a,l}^{(\text{V},v)}(t) - \frac{S_{a,l}^{(\text{PI},v)}(t)}{N}\sum_{m\in M}\lambda_{a,l}^{(\text{PI},m,v)}(t)\\ &= \alpha_a^{(v)} f_S^{(\text{PI},v)} S_{a,l}^{(\text{V},v)}(t) - \frac{S_{a,l}^{(\text{PI},v)}(t)}{N}\sum_{m\in M}g(m,v)\lambda_{a,l}^{(m)}(t).\end{aligned} \tag{19c}$$

The number of unimmunized individuals changes because fractions of vaccinated susceptibles completely fail to be immunized after vaccination (with one of the vaccines) and due to the forces of infection. Their numbers changes according to

$$\frac{dS_{a,l}^{(\mathrm{NI})}(t)}{dt} = \sum_{v \in V} \alpha_a^{(v)} f_S^{(\mathrm{NI},v)} S_{a,l}^{(\mathrm{V},v)}(t) - \frac{S_{a,l}^{(\mathrm{NI})}(t)}{N} \sum_{m \in M} \lambda_{a,l}^{(m)}(t). \tag{19d}$$

Finally, a fraction of individuals will be completely immunized by vaccine $v$ against at least one viral variant. However, they might be susceptible to other viral variants, so that their number changes according to

$$\frac{dR_{a,l}^{(\mathrm{Im},v)}(t)}{dt} = \alpha_a^{(v)} f_S^{(\mathrm{Im},v)} S_{a,l}^{(\mathrm{V},v)}(t) - \frac{R_{a,l}^{(\mathrm{Im},v)}(t)}{N} \sum_{m \in M} h(m,v) \lambda_{a,l}^{(m)}(t). \tag{19e}$$

### Dynamics of latent-infected individuals

Susceptibles transit to the latent phase upon infection. Latent-infected individuals progress through $n_E^{(a,m)}$ sub-states at rate $\varepsilon_{a,m}$. The number of sub-states and the rate depend on the infecting variant and the age group of the infected individual. Unvaccinated latent-infected individuals will still get vaccinated. Hence, the numbers of individuals latently infected with variant $m$ in age group $a$ and location $l$ changes according to

$$\frac{dE_{1,a,l}^{(\mathrm{U},m)}(t)}{dt} = \lambda_{a,l}^{(m)}(t) \frac{S_{a,l}^{(\mathrm{U})}(t)}{N} - \Big(\varepsilon_{a,m} + \sum_{v \in V} \nu_{a,l}^{(v)}\Big) E_{1,a,l}^{(\mathrm{U},m)}(t), \tag{20a}$$

and

$$\frac{dE_{k,a,l}^{(\mathrm{U},m)}(t)}{dt} = \varepsilon_{a,m} E_{k-1,a,l}^{(\mathrm{U},m)}(t) - \Big(\varepsilon_{a,m} + \sum_{v \in V} \nu_{a,l}^{(v)}\Big) E_{k,a,l}^{(\mathrm{U},m)}(t), \tag{20b}$$

where (here and in the following) $k = 2, \ldots, n_E^{(a,m)}$, $m \in M$, and $v \in V$. The numbers of latent infected individuals vaccinated with $v$ with pending vaccination outcome changes according to

$$\frac{dE_{1,a,l}^{(\mathrm{V},m,v)}(t)}{dt} = \lambda_{a,l}^{(m)}(t) \frac{S_{a,l}^{(\mathrm{V},v)}(t)}{N} + \nu_{a,l}^{(v)} E_{1,a,l}^{(\mathrm{U},m)}(t) - \big(\alpha_a^{(v)} + \varepsilon_{a,m}\big) E_{1,a,l}^{(\mathrm{V},m,v)}(t), \tag{20c}$$

and

$$\frac{dE_{k,a,l}^{(\mathrm{V},m,v)}(t)}{dt} = \varepsilon_{a,m} E_{k-1,a,l}^{(\mathrm{V},m,v)}(t) + \nu_{a,l}^{(v)} E_{k,a,l}^{(\mathrm{U},m)}(t) - \big(\alpha_a^{(v)} + \varepsilon_{a,m}\big) E_{k,a,l}^{(\mathrm{V},m,v)}(t). \tag{20d}$$

Individuals vaccinated with $v$ might become latently infected by a variant $m$, if $v$ only failed to completely immunize against this variant (either because $v$ immunized completely against another viral variant, or $v$ did not or only partially immunized against all variants). Moreover, the partial immunization of $v$ might manifest during the latent phase. Hence, the numbers of partially-immune latent infected individuals changes according to

$$\begin{aligned}\frac{dE_{1,a,l}^{(\mathrm{PI},m,v)}(t)}{dt} =& \lambda_{a,l}^{(\mathrm{PI},m,v)}(t)\frac{S_{a,l}^{(\mathrm{PI},v)}(t)}{N} + \lambda_{a,l}^{(\mathrm{Im},m,v)}(t)\frac{R_{a,l}^{(\mathrm{Im},v)}(t)}{N} \\ &+ \alpha_a^{(v)} f_E^{(\mathrm{PI},v)} E_{1,a,l}^{(\mathrm{V},m,v)}(t) - \varepsilon_{a,m} E_{1,a,l}^{(\mathrm{PI},m,v)}(t),\end{aligned} \tag{20e}$$

and

$$\frac{dE_{k,a,l}^{(\mathrm{PI},m,v)}(t)}{dt} = \varepsilon_{a,m} E_{k-1,a,l}^{(\mathrm{PI},m,v)}(t) + \alpha_a^{(v)} f_E^{(\mathrm{PI},v)} E_{k,a,l}^{(\mathrm{V},m,v)}(t) - \varepsilon_{a,m} E_{k,a,l}^{(\mathrm{PI},m,v)}(t). \tag{20f}$$

If infected, individuals that are non-immunized progress to the latent phase. So do infected vaccinated individuals if the vaccine failed to immunize. Hence, the dynamics of non-immunized latently infected individuals become

$$\frac{dE_{1,a,l}^{(\mathrm{NI},m)}(t)}{dt} = \lambda_{a,l}^{(m)}(t)\frac{S_{a,l}^{(\mathrm{NI})}(t)}{N} - \varepsilon_{a,m} E_{1,a,l}^{(\mathrm{NI},m)}(t) + \sum_{v\in V} \alpha_a^{(v)} f_E^{(\mathrm{NI},v)} E_{1,a,l}^{(\mathrm{V},m,v)}(t), \tag{20g}$$

and

$$\frac{dE_{k,a,l}^{(\mathrm{NI},m)}(t)}{dt} = \varepsilon_{a,m} E_{k-1,a,l}^{(\mathrm{NI},m)}(t) - \varepsilon_{a,m} E_{k,a,l}^{(\mathrm{NI},m)}(t) + \sum_{v\in V} \alpha_a^{(v)} f_E^{(\mathrm{NI},v)} E_{k,a,l}^{(\mathrm{V},m,v)}(t). \tag{20h}$$

#### Dynamics of prodromal individuals

The dynamics of the prodromal states are similar to those of the latent states, namely, (with $k = 2, \ldots, n_P^{(a,m)}$, $m \in M$ and $v \in V$):

$$
\begin{aligned}
\frac{dP_{1,a,l}^{(\mathrm{U},m)}(t)}{dt} =& \varepsilon_{a,m} E_{n_E,a,l}^{(\mathrm{U},m)}(t) - \big(\varphi_{a,m} + \sum_{v \in V} \nu_{a,l}^{(v)}\big) P_{1,a,l}^{(\mathrm{U},m)}(t), && \text{(21a)}\\
\frac{dP_{k,a,l}^{(\mathrm{U},m)}(t)}{dt} =& \varphi_{a,m} P_{k-1,a,l}^{(\mathrm{U},m)}(t) - \big(\varphi_{a,m} + \sum_{v \in V} \nu_{a,l}^{(v)}\big) P_{k,a,l}^{(\mathrm{U},m)}(t), && \text{(21b)}\\
\frac{dP_{1,a,l}^{(\mathrm{V},m,v)}(t)}{dt} =& \varepsilon_{a,m} E_{n_E,a,l}^{(\mathrm{V},m,v)}(t) + \nu_{a,l}^{(v)} P_{1,a,l}^{(\mathrm{U},m)}(t) - \big(\varphi_{a,m} + \alpha_a^{(v)}\big) P_{1,a,l}^{(\mathrm{V},m,v)}(t), && \text{(21c)}\\
\frac{dP_{k,a,l}^{(\mathrm{V},m,v)}(t)}{dt} =& \varphi_{a,m} P_{k-1,a,l}^{(\mathrm{V},m,v)}(t) + \nu_{a,l}^{(v)} P_{k,a,l}^{(\mathrm{U},m)}(t) - \big(\varphi_{a,m} + \alpha_a^{(v)}\big) P_{k,a,l}^{(\mathrm{V},m,v)}(t), && \text{(21d)}\\
\frac{dP_{1,a,l}^{(\mathrm{PI},m,v)}(t)}{dt} =& \varepsilon_{a,m} E_{n_E,a,l}^{(\mathrm{PI},m,v)}(t) + \alpha_a^{(v)} f_P^{(\mathrm{PI},v)} P_{1,a,l}^{(\mathrm{V},m,v)}(t) - \varphi_{a,m} P_{1,a,l}^{(\mathrm{PI},m,v)}(t), && \text{(21e)}\\
\frac{dP_{k,a,l}^{(\mathrm{PI},m,v)}(t)}{dt} =& \varphi_{a,m} P_{k-1,a,l}^{(\mathrm{PI},m,v)}(t) + \alpha_a^{(v)} f_P^{(\mathrm{PI},v)} P_{k,a,l}^{(\mathrm{V},m,v)}(t) - \varphi_{a,m} P_{k,a,l}^{(\mathrm{PI},m,v)}(t), && \text{(21f)}\\
\frac{dP_{1,a,l}^{(\mathrm{NI},m)}(t)}{dt} =& \varepsilon_{a,m} E_{n_E,a,l}^{(\mathrm{NI},m)}(t) + \sum_{v \in V} \alpha_a^{(v)} f_P^{(\mathrm{NI},v)} P_{1,a,l}^{(\mathrm{V},m,v)}(t) - \varphi_{a,m} P_{1,a,l}^{(\mathrm{NI},m)}(t), && \text{(21g)}\\
\frac{dP_{k,a,l}^{(\mathrm{NI},m)}(t)}{dt} =& \varphi_{a,m} P_{k-1,a,l}^{(\mathrm{NI},m)}(t) + \sum_{v \in V} \alpha_a^{(v)} f_P^{(\mathrm{NI},v)} P_{k,a,l}^{(\mathrm{V},m,v)}(t) - \varphi_{a,m} P_{k,a,l}^{(\mathrm{NI},m)}(t). && \text{(21h)}
\end{aligned}
$$

#### Dynamics of fully-infectious individuals

The dynamics for the fully-infectious states are similar to those of the prodromal states. However, it is assumed that a vaccine will not have an effect if individuals are vaccinated during this state, so that vaccination can be ignored. Hence, for

$k=2,\dots,n_I^{(a,m)}$, $m\in M$ and $v\in V$ we have

$$\frac{dI_{1,a,l}^{(\mathrm{U},m)}(t)}{dt} = \varphi_{a,m}P_{n_P,a,l}^{(\mathrm{U},m)}(t) - \gamma_{a,m}I_{1,a,l}^{(\mathrm{U},m)}(t), \tag{22a}$$

$$\frac{dI_{k,a,l}^{(\mathrm{U},m)}(t)}{dt} = \gamma_{a,m}I_{k-1,a,l}^{(\mathrm{U},m)}(t) - \gamma_{a,m}I_{k,a,l}^{(\mathrm{U},m)}(t), \tag{22b}$$

$$\frac{dI_{1,a,l}^{(\mathrm{V},m,v)}(t)}{dt} = \varphi_{a,m}P_{n_P,a,l}^{(\mathrm{V},m,v)}(t) - \big(\gamma_{a,m}+\alpha_a^{(v)}\big)I_{1,a,l}^{(\mathrm{V},m,v)}(t), \tag{22c}$$

$$\frac{dI_{k,a,l}^{(\mathrm{V},m,v)}(t)}{dt} = \gamma_{a,m}I_{k-1,a,l}^{(\mathrm{V},m,v)}(t) - \big(\gamma_{a,m}+\alpha_a^{(v)}\big)I_{k,a,l}^{(\mathrm{V},m,v)}(t), \tag{22d}$$

$$\frac{dI_{1,a,l}^{(\mathrm{PI},m,v)}(t)}{dt} = \varphi_{a,m}P_{n_P,a,l}^{(\mathrm{PI},m,v)}(t) + \alpha_a^{(v)}f_I^{(\mathrm{PI},v)}I_{1,a,l}^{(\mathrm{V},m,v)}(t) - \gamma_{a,m}I_{1,a,l}^{(\mathrm{PI},m,v)}(t), \tag{22e}$$

$$\frac{dI_{k,a,l}^{(\mathrm{PI},m,v)}(t)}{dt} = \gamma_{a,m}I_{k-1,a,l}^{(\mathrm{PI},m,v)}(t) + \alpha_a^{(v)}f_I^{(\mathrm{PI},v)}I_{k,a,l}^{(\mathrm{V},m,v)}(t) - \gamma_{a,m}I_{k,a,l}^{(\mathrm{PI},m,v)}(t), \tag{22f}$$

$$\frac{dI_{1,a,l}^{(\mathrm{NI},m)}(t)}{dt} = \varphi_{a,m}P_{n_P,a,l}^{(\mathrm{NI},m)}(t) + \sum_{v\in V}\alpha_a^{(v)}f_I^{(\mathrm{NI},v)}I_{1,a,l}^{(\mathrm{V},m,v)}(t) - \gamma_{a,m}I_{1,a,l}^{(\mathrm{NI},m)}(t), \tag{22g}$$

$$\frac{dI_{k,a,l}^{(\mathrm{NI},m)}(t)}{dt} = \gamma_{a,m}I_{k-1,a,l}^{(\mathrm{NI},m)}(t) + \sum_{v\in V}\alpha_a^{(v)}f_I^{(\mathrm{NI},v)}I_{k,a,l}^{(\mathrm{V},m,v)}(t) - \gamma_{a,m}I_{k,a,l}^{(\mathrm{NI},m)}(t). \tag{22h}$$

### Dynamics of late-infectious individuals

The dynamics of the late-infectious states are similar to those of the fully-infectious states with obvious modifications. Namely, for $k=2,\dots,n_L^{(a,m)}$, $m\in M$ and $v\in V$ one obtains

$$\frac{dL_{1,a,l}^{(\mathrm{U},m)}(t)}{dt} = \gamma_{a,m}I_{n_I,a,l}^{(\mathrm{U},m)}(t) - \delta_{a,m}L_{1,a,l}^{(\mathrm{U},m)}(t), \tag{23a}$$

$$\frac{dL_{k,a,l}^{(\mathrm{U},m)}(t)}{dt} = \delta_{a,m}L_{k-1,a,l}^{(\mathrm{U},m)}(t) - \delta_{a,m}L_{k,a,l}^{(\mathrm{U},m)}(t), \tag{23b}$$

$$\frac{dL_{1,a,l}^{(\mathrm{V},m,v)}(t)}{dt} = \gamma_{a,m}I_{n_I,a,l}^{(\mathrm{V},m,v)}(t) - \big(\delta_{a,m}+\alpha_a^{(v)}\big)L_{1,a,l}^{(\mathrm{V},m,v)}(t), \tag{23c}$$

$$\frac{dL_{k,a,l}^{(\mathrm{V},m,v)}(t)}{dt} = \delta_{a,m}L_{k-1,a,l}^{(\mathrm{V},m,v)}(t) - \big(\delta_{a,m}+\alpha_a^{(v)}\big)L_{k,a,l}^{(\mathrm{V},m,v)}(t), \tag{23d}$$

$$\frac{dL_{1,a,l}^{(\mathrm{PI},m,v)}(t)}{dt} = \gamma_{a,m}I_{n_I,a,l}^{(\mathrm{PI},m,v)}(t) + \alpha_a^{(v)}f_L^{(\mathrm{PI},v)}I_{1,a,l}^{(\mathrm{V},m,v)}(t) - \delta_{a,m}L_{1,a,l}^{(\mathrm{PI},m,v)}(t), \tag{23e}$$

$$\frac{dL_{k,a,l}^{(\mathrm{PI},m,v)}(t)}{dt} = \delta_{a,m}L_{k-1,a,l}^{(\mathrm{PI},m,v)}(t) + \alpha_a^{(v)}f_L^{(\mathrm{PI},v)}L_{k,a,l}^{(\mathrm{V},m,v)}(t) - \delta_{a,m}L_{k,a,l}^{(\mathrm{PI},m,v)}(t), \tag{23f}$$

$$\frac{dL_{1,a,l}^{(\mathrm{NI},m)}(t)}{dt} = \gamma_{a,m}I_{n_I,a,l}^{(\mathrm{NI},m)}(t) + \sum_{v\in V}\alpha_a^{(v)}f_L^{(\mathrm{NI},v)}L_{1,a,l}^{(\mathrm{V},m,v)}(t) - \delta_{a,m}L_{1,a,l}^{(\mathrm{NI},m)}(t), \tag{23g}$$

$$\frac{dL_{k,a,l}^{(\mathrm{NI},m)}(t)}{dt} = \delta_{a,m}L_{k-1,a,l}^{(\mathrm{NI},m)}(t) + \sum_{v\in V}\alpha_a^{(v)}f_L^{(\mathrm{NI},v)}L_{k,a,l}^{(\mathrm{V},m,v)}(t) - \delta_{a,m}L_{k,a,l}^{(\mathrm{NI},m)}(t). \tag{23h}$$

### Dynamics of recovered individuals

Following the last late-infectious state, asymptomatic infections lead to recovery. So does the fraction of symptomatic infections that are not lethal. Moreover, if the effect of the vaccine manifests during any state of the infection, it can lead to early recovery. Hence, the number of recovered individuals in age group $a$ in location $l$ that were infected with variant $m$ changes according to

$$\begin{aligned}\frac{dR_{a,l}^{(\mathrm{Inf},m)}(t)}{dt} =& \sum_{v\in V}\alpha_a^{(v)}\Bigg(f_E^{(\mathrm{Im},v)}\sum_{k=1}^{n_E^{(a,m)}} E_{k,a,l}^{(\mathrm{V},m,v)}(t) + f_P^{(\mathrm{Im},v)}\sum_{k=1}^{n_P^{(a,m)}} P_{k,a,l}^{(\mathrm{V},m,v)}(t)\\ &+ f_I^{(\mathrm{Im},v)}\sum_{k=1}^{n_I^{(a,m)}} I_{k,a,l}^{(\mathrm{V},m,v)}(t) + f_L^{(\mathrm{Im},v)}\sum_{k=1}^{n_L^{(a,m)}} L_{k,a,l}^{(\mathrm{V},m,v)}(t)\Bigg)\\ &+\delta_{a,m}\left(1-f_{\mathrm{Sick}}^{(a,m)}f_{\mathrm{Dead}}^{(a,m)}\right)\left(L_{n_L,a,l}^{(\mathrm{U},m)}(t)+\sum_{v\in V}L_{n_L,a,l}^{(\mathrm{V},m,v)}(t)+L_{n_L,a,l}^{(\mathrm{NI},m)}(t)\right)\\ &+\delta_{a,m}\sum_{v\in V}\left(1-f_{\mathrm{Sick}}^{(\mathrm{PI},a,m,v)}f_{\mathrm{Dead}}^{(\mathrm{PI},a,m,v)}\right)L_{n_L,a,l}^{(\mathrm{PI},m,v)}(t).\end{aligned} \tag{24}$$

### Dead individuals

A fraction of symptomatic infections is lethal. Death occurs after the last late-infectious state. The number of deaths changes according to

$$\begin{aligned}\frac{dD_{a,l}^{(m)}(t)}{dt} =& \delta_{a,m}f_{\mathrm{Sick}}^{(a,m)}f_{\mathrm{Dead}}^{(a,m)}\left(L_{n_L,a,l}^{(\mathrm{U},m)}(t)+\sum_{v\in V}L_{n_L,a,l}^{(\mathrm{V},m,v)}(t)+L_{n_L,a,l}^{(\mathrm{NI},m)}(t)\right)\\ &+\delta_{a,m}\sum_{v\in V}f_{\mathrm{Sick}}^{(\mathrm{PI},a,m,v)}f_{\mathrm{Dead}}^{(\mathrm{PI},a,m,v)}L_{n_L,a,l}^{(\mathrm{PI},m,v)}(t).\end{aligned} \tag{25}$$

#### Mathematical description of the demographic mixing matrix

Let $N_{a,l}$ be the number of individuals in age group $a$ and location $l$. Clearly, $N = \sum_{a=1}^{s}\sum_{l=1}^{r} N_{a,l}$. Moreover, let $n_{a,l}$ be the average number of contacts, which an individual in age group $a$ and location $l$ has. These contacts are distributed across the whole population. Let $c_{a',l'}^{(a,l)}$ be the proportion of contacts that individuals in age group $a$ and location $l$ have with individuals in age group $a'$ and location $l'$. This is a probability distribution, i.e., $\sum_{a'=1}^{s}\sum_{l'=1}^{r} c_{a',l'}^{(a,l)} = 1$. Hence, the average number of contacts (encounters) in age group $a$ and location $l$ with individuals in age group $a'$ and location $l'$ is given by $e_{a',l'}^{(a,l)} = c_{a',l'}^{(a,l)} n_{a,l} N_{a,l}$. Since contacts are bidirectional, the relation $e_{a',l'}^{(a,l)} = e_{a,l}^{(a',l')}$ must hold, i.e.,

$$c_{a',l'}^{(a,l)} n_{a,l} N_{a,l} = c_{a,l}^{(a',l')} n_{a',l'} N_{a',l'}. \tag{26a}$$

Assuming random encounters, the probability of contacts between age group $a$ in location $l$ with individuals from age group $a'$ in location $l'$, would be

$$r_{a',l'}^{(a,l)} := r_{a,l}^{(a',l')} := \frac{N_{a,l}}{N}\frac{N_{a',l'}}{N}. \tag{26b}$$

The matrix consisting of the encounters $e^{(a,l)}_{a',l'}$ is referred to as the encounter matrix $E_{\text{Tot}}$. The mixing matrix $X$, consisting of entries $x^{(a,l)}_{a',l'}$, adjusts random encounters in such a way that the true contact behavior emerges, i.e.,

$$x^{(a,l)}_{a',l'} := e^{(a,l)}_{a',l'}/r^{(a,l)}_{a',l'} = \frac{c^{(a,l)}_{a',l'} n_{a,l} N^2}{N_{a',l'}}. \tag{26c}$$

##### General contact reduction

General contact reduction is sustained in a time-dependent fashion. At each time point, the amount of contacts being reduced depends on the characteristics of the interactions between sub-populations. This changes the matrix of encounters $E_{\text{Tot}}$ to $\tilde{E}_{\text{Tot}}(t)$.

For instance, let $p^{(a,l,a',l')}_{\text{Cont}}(t) = p^{(a',l',a,l)}_{\text{Cont}}(t)$ be the fraction of contacts that is reduced between age group $a$ in location $l$ and age group $a'$ in location $l'$ at time $t$. The actual amounts of these reductions depend on the type of interventions being sustained. Then, the matrix $\tilde{E}_{\text{Tot}}(t)$ has entries

$$\tilde{e}^{(a,l)}_{a',l'}(t) = e^{(a,l)}_{a',l'}\big(1 - p^{(a,l,a',l')}_{\text{Cont}}(t)\big).$$

For all simulations described throughout this article, more refined contact reductions are assumed. In particular, considering just one location, the matrix $E_{\text{Tot}}$ is partitioned into contacts at home ($E^{(\text{Home})}_{\text{Tot}}$), school ($E^{(\text{School})}_{\text{Tot}}$), work, ($E^{(\text{Work})}_{\text{Tot}}$) and other contacts ($E^{(\text{Others})}_{\text{Tot}}$) describing leisure time. Contacts at home and leisure time are further subdivided into contacts between the risk group of elderly individuals ($E^{(\text{Home})}_{\text{Tot}}$ and $E^{(\text{Home, Old})}_{\text{Tot}}$; $E^{(\text{Others})}_{\text{Tot}}$ and $E^{(\text{Others, Old})}_{\text{Tot}}$). Hence,

$$E_{\text{Tot}} = E^{(\text{Home})}_{\text{Tot}} + E^{(\text{Home, Old})}_{\text{Tot}} + E^{(\text{School})}_{\text{Tot}} + E^{(\text{Work})}_{\text{Tot}} + E^{(\text{Others})}_{\text{Tot}} + E^{(\text{Others, Old})}_{\text{Tot}}.$$

Contact reductions apply then to different categories of contacts, i.e.,

$$\begin{aligned}\tilde{E}_{\text{Tot}}(t) =& \Big(1 - p^{(\text{Home})}_{\text{Cont}}(t)\Big) E^{(\text{Home})}_{\text{Tot}} + \Big(1 - p^{(\text{Home, Old})}_{\text{Cont}}(t)\Big) E^{(\text{Home, Old})}_{\text{Tot}} \\ &+ \Big(1 - p^{(\text{School})}_{\text{Cont}}(t)\Big) E^{(\text{School})}_{\text{Tot}} + \Big(1 - p^{(\text{Work})}_{\text{Cont}}(t)\Big) E^{(\text{Work})}_{\text{Tot}} \\ &+ \Big(1 - p^{(\text{Others})}_{\text{Cont}}(t)\Big) E^{(\text{Others})}_{\text{Tot}} + \Big(1 - p^{(\text{Others, Old})}_{\text{Cont}}(t)\Big) E^{(\text{Others, Old})}_{\text{Tot}}.\end{aligned}$$

Here, contact reductions are also age dependent as the stratification of contacts into the different categories (Home, School, Work, Others) is age-dependent. The advantage of such a parameterization is that it is easily possible to reflect COVID-19 management policies. For more than one location, similar contact reductions need to be specified per location and between locations.

Once the time-dependent matrix of total contacts $\tilde{E}_{\text{Tot}}(t)$ is specified, the entries of the mixing matrix $X$ become a function of time defined by

$$x^{(a,l)}_{a',l'}(t) := \tilde{e}^{(a,l)}_{a',l'}(t)/r^{(a,l)}_{a',l'}. \tag{27}$$

##### Incidence-based general contact reduction

General contact reduction are time dependent in general. As a special case of time-dependence they can be incidence-based during some time periods, as implemented for instance in Germany [5]. Importantly, incidence-based interventions do not distinguish between the viral variants. The "overall" incidence rate at location $l$ is defined as

$$\iota_l(t) := \sum_{a=1}^{s} \sum_{m\in M} \sum_{v\in V} \Bigg[ \lambda_{a,l}^{(m)}(t) \frac{S_{a,l}^{(\mathrm{U})}(t) + S_{a,l}^{(\mathrm{V},v)}(t) + S_{a,l}^{(\mathrm{NI})}(t)}{N} + \lambda_{a,l}^{(\mathrm{PI},m,v)}(t) \frac{S_{a,l}^{(\mathrm{PI},v)}(t)}{N} + \lambda_{a,l}^{(\mathrm{Im},m,v)}(t) \frac{R_{a,l}^{(\mathrm{Im},v)}(t)}{N} \Bigg]. \tag{28}$$

The average incidence over $d$-time units (in practice this will be chosen as the 7-day-average incidence) in location $l$ is defined to be

$$i_l(t) := \frac{1}{d} \int_{t-d}^{t} \iota^{(l)}(s) ds. \tag{29}$$

The $d$-days incidence is defined as the new infections in a location over $d$-days per 100 000 individuals, i.e., as

$$i_l(t) \frac{10^5 d}{N_l}. \tag{30}$$

In practice, the 7-days incidence is used.

The contact reduction between individuals from age groups $a$ and $a'$ in locations $l$ and $l'$ are defined as

$$p_{\mathrm{Cont}}^{(a,l,a',l')}(t) = p_{\mathrm{Tr},u,w}^{(a,l,a',l')} \text{ if } i_{\mathrm{Tr},u-1}^{(l)} \leq i_l(t) < i_{\mathrm{Tr},u}^{(l)} \text{ and } i_{\mathrm{Tr},w-1}^{(l')} \leq i_{l'}(t) < i_{\mathrm{Tr},w}^{(l')}, \tag{31}$$

where $0 = i_{\mathrm{Tr},0}^{(l)} < \ldots < i_{\mathrm{Tr},A}^{(l)} = \infty$ are the incidence thresholds in location $l$. These are the threshold values for the average incidence at which the imposed general contact reductions in location $l$ change. Changes in contact reductions in location $l$ has implications on location $l'$, because mobility between these locations can be affected by contact reductions in any of the two locations.

#### The basic reproduction number and the next generation matrix

The basic reproduction number $R_0$ is defined as the average number of infections caused by an infected individual in a completely susceptible population. This definition holds only for homogeneous populations, not for heterogeneous subdivided population. In the latter case it is rather the average number of infections caused by an average infected individual [4] in a susceptible population without any disease-control interventions. As such $R_0$ is derived as the maximum eigenvalue of the next generation matrix (NGM). The presentation follows [3].

To derive the NGM, first the reduced system of ODEs assuming no control interventions, in which the whole population is susceptible, needs to be linearized. The reduced system consists only of those differential equations from the original system that describe the states of infected individuals, which are relevant in the absence of interventions. Here, this implies the absence of vaccinations, cases isolation, or general contact reduction. Further, only the presence of a single viral variant has to be considered. Moreover, the distinction between unvaccinable and individuals waiting to be vaccinated is irrelevant. In the present case the reduced system becomes,

$$\frac{dE_{1,a,l}(t)}{dt} = \lambda_{a,l}(t)\frac{S_{a,l}(t)}{N} - \varepsilon_a E_{1,a,l}(t), \tag{32a}$$

$$\frac{dE_{k,a,l}(t)}{dt} = \varepsilon_a E_{k-1,a,l}(t) - \varepsilon_a E_{k,a,l}(t) \qquad \text{for} \quad 2 \le k \le n_E^{(a)}, \tag{32b}$$

$$\frac{dP_{1,a,l}(t)}{dt} = \varepsilon_a E_{n_E,a,l}(t) - \varphi_a P_{1,a,l}(t), \tag{32c}$$

$$\frac{dP_{k,a,l}(t)}{dt} = \varphi_a P_{k-1,a,l}(t) - \varphi_a P_{k,a,l}(t) \qquad \text{for} \quad 2 \le k \le n_P^{(a)}, \tag{32d}$$

$$\frac{dI_{1,a,l}(t)}{dt} = \varphi_a P_{n_P,a,l}(t) - \gamma_a I_{1,a,l}(t), \tag{32e}$$

$$\frac{dI_{k,a,l}(t)}{dt} = \gamma_a I_{k-1,a,l}(t) - \gamma_a I_{k,a,l}(t) \qquad \text{for} \quad 2 \le k \le n_I^{(a)}, \tag{32f}$$

$$\frac{dL_{1,a,l}(t)}{dt} = \gamma_a I_{n_I,a,l}(t) - \delta_a L_{1,a,l}(t), \tag{32g}$$

$$\frac{dL_{k,a,l}(t)}{dt} = \delta_a L_{k-1,a,l}(t) - \delta_a L_{k,a,l}(t) \qquad \text{for} \quad 2 \le k \le n_L^{(a)}. \tag{32h}$$

All states of the reduced system are denoted by vector $\boldsymbol{x}$. The initial state, in which all individuals are susceptible is denoted by $\boldsymbol{x}_0$. The Jacobian of the reduced system is split into one matrix describing transmission and one matrix describing transitions between infected states. For this purpose, vector-valued functions $F(\boldsymbol{x})$ and $V(\boldsymbol{x})$, describing transmission and transitions in the reduced system are defined, whose sum of Jacobian matrices, $\frac{\partial F}{\partial \boldsymbol{x}}$ and $\frac{\partial V}{\partial \boldsymbol{x}}$, equals the Jacobian of the reduced system. Once, these Jacobians are derived, the NGM is calculated as

$$G := -\left[\frac{\partial F}{\partial \boldsymbol{x}}(\boldsymbol{x}_0)\right]\left[\frac{\partial V}{\partial \boldsymbol{x}}(\boldsymbol{x}_0)\right]^{-1}. \tag{33}$$

The basic reproduction number $R_0$ is defined as the spectral radius of the matrix $G$ (cf. [4]). In mathematical terms

$$R_0 := \rho(G) := \max_i |\lambda_i(G)|, \tag{34}$$

where $\lambda_i(G)$ denote the eigenvalues of $G$. Note that $\rho(G)$ is a function of $\bar{R}_0^{(\mathrm{adj})}$. It has to be chosen such that

$$\bar{R}_0 = \frac{\max_i |\lambda_i(G)|}{1 + A\cos\left(-2\pi\frac{t_{R_{0_{\max}}}}{365}\right)}$$

holds.

Here, the non-zero entries of the function $F(\boldsymbol{x}(t))$, indexed by compartment names, are

$$F_{E_{1,a,l}}(t) = \lambda_{a,l}(t)\frac{S_{a,l}(t)}{N} \quad \text{for} \quad a = 1, \dots, s \quad \text{and} \quad l = 1, \dots, r.$$

Thus, the non-zero entries of the partial derivatives $\dfrac{\partial F}{\partial \boldsymbol{x}}$ are for $a$, $a' = 1, \dots, s$ and $l$, $l' = 1, \dots, r$,

$$
\begin{aligned}
\frac{\partial F_{E_{1,a,l}}(t)}{\partial P_{k,a',l'}} &= x^{(a,l)}_{a',l'} \beta^{(a')}_P(t) \frac{S_{a,l}(t)}{N} && \text{for} \quad 1 \le k \le n^{(a)}_P, && (35a)\\
\frac{\partial F_{E_{1,a,l}}(t)}{\partial I_{k,a',l'}} &= x^{(a,l)}_{a',l'}(t) \beta^{(a')}_I(t) \frac{S_{a,l}(t)}{N} && \text{for} \quad 1 \le k \le n^{(a)}_I, && (35b)\\
\frac{\partial F_{E_{1,a,l}}(t)}{\partial L_{k,a',l'}} &= x^{(a,l)}_{a',l'}(t) \beta^{(a')}_L(t) \frac{S_{a,l}(t)}{N} && \text{for} \quad 1 \le k \le n^{(a)}_L. && (35c)
\end{aligned}
$$

The non-zero components of the function $V(\boldsymbol{x}(t))$ are

$$
\begin{aligned}
V_{E_{1,a,l}}(t) &= -\varepsilon_a E_{1,a,l}(t), && && (36a)\\
V_{E_{k,a,l}}(t) &= \varepsilon_a E_{k-1,a,l}(t) - \varepsilon_a E_{k,a,l}(t) && \text{for} \quad 2 \le k \le n^{(a)}_E, && (36b)\\
V_{P_{1,a,l}}(t) &= \varepsilon_a E_{n_E,a,l}(t) - \varphi_a P_{1,a,l}(t), && && (36c)\\
V_{P_{k,a,l}}(t) &= \varphi_a P_{k-1,a,l}(t) - \varphi_a P_{k,a,l}(t) && \text{for} \quad 2 \le k \le n^{(a)}_P, && (36d)\\
V_{I_{1,a,l}}(t) &= \varphi_a P_{n_P,a,l}(t) - \gamma_a I_{1,a,l}(t), && && (36e)\\
V_{I_{k,a,l}}(t) &= \gamma_a I_{k-1,a,l}(t) - \gamma_a I_{k,a,l}(t) && \text{for} \quad 2 \le k \le n^{(a)}_I, && (36f)\\
V_{L_{1,a,l}}(t) &= \gamma_a I_{n_I,a,l}(t) - \delta_a L_{1,a,l}(t), && && (36g)\\
V_{L_{k,a,l}}(t) &= \delta_a L_{k-1,a,l}(t) - \delta_a L_{k,a,l}(t) && \text{for} \quad 2 \le k \le n^{(a)}_L. && (36h)
\end{aligned}
$$

The Jacobian of (36) is straightforward. Hence, (33) can be easily derived from (35) and the derivatives of (36). Let

$$
\tilde{G} := \frac{1}{\bar{R}^{(\mathrm{adj})}_0 \left(1 + A \cos\left(-2\pi \frac{t_{R_{0\max}}}{365}\right)\right)} G, \tag{37}
$$

which is independent of $\bar{R}^{(\mathrm{adj})}_0$. Then

$$
\bar{R}^{(\mathrm{adj})}_0 = \frac{\bar{R}_0}{\max\limits_i |\lambda_i(\tilde{G})|}. \tag{38}
$$

The adjusted basic reproduction number $\bar{R}^{(m,\mathrm{adj})}_0$ of viral variant $m$ is then given by substituting $\bar{R}^{(m)}_0$ for $\bar{R}_0$ in (38).

#### Weather adjustment

Seasonal fluctuations in the basic reproduction number are crucial to obtain realistic dynamics. Clearly, (7) assumes harmonic oscillation of $R^{(m)}_0(t)$. This implicitly assume that weather is periodic in one-year periods. This might be different in some tropical and subtropical areas, characterized for instance by two wet and two dry seasons each year. Moreover, weather is not the only factor influencing the basic reproduction number. Factors correlating with weather also influence the contact behavior, e.g., although in summer high UV radiation would decrease transmissibility, it might increase if people spend their time on crowded beaches. Therefore, (7) needs to be adjusted as appropriate for the application. One important instance are unusually warm or cold months during the year, that act like a shift in weather. To adjust weather by a shift $\Delta$, (7) needs to be replaced by

$$R_0^{(m)}(t) := \bar{R}_0^{(m)}\left(1 + A\cos\left(2\pi\frac{t - \Delta - t_{R_{0_{\max}}}}{365}\right)\right). \tag{39}$$

This means that weather at time $t$ is actually behaving like weather at time $t - \Delta$.

5. Bundesanzeiger Verlag GmbH. Bundesgesetzblatt Jahrgang 2021 Teil I; 2021. Nr. 18.
