## Supplementary material for "Predicting the impact of COVID-19 vaccination campaigns – a flexible age-dependent, spatially-stratified predictive model, accounting for multiple viral variants and vaccines": S1 Simulation results for Saxony and Schleswig-Holstein

### S1 Simulations for Saxony and Schleswig-Holstein

In the main text the model was parameterized to reflect the COVID-19 epidemic in Germany. Although this accurately reflected the situation in Germany, it is Europe's most populated and third largest country governed by a federalist system. Epidemic management in Germany was partly under the legislation of its provinces, which are diverse in size and culture. When attempting to model Germany on a more fine-grained scale, the provinces can be modeled as separate locations. This would require information on the contact reductions for each of Germany's provinces. However, this is beyond the scope of this article. Therefore, we selected two provinces, which differ in several important characteristics and modeled them together, namely Saxony and Schleswig-Holstein.

Saxony is a province in former East Germany bordering the Czech Republic and Poland, with a relatively high proportion of elderly citizens. Moreover, in Saxony vaccination hesitancy is widespread, in fact Saxony currently has the lowest vaccination coverage among all provinces (less than 60%) [1, 2]. Despite this fact, COVID-19 incidence remained low in Saxony, when incidences were already raising in other provinces in which a much larger part of the population was vaccinated. Schleswig-Holstein is one of those. It is Germany's northmost province bordering Denmark, in between the North and the East Sea. It ranks among the provinces with the highest vaccination coverages (almost 75%) [1]. Since Germany has a sophisticated highway and railways system, mobility exchange between these two provinces occurs.

#### Parameter choices

##### Population and contact behavior

The population sizes $N_1 = 4\,085\,344$ for Saxony and $N_2 = 2\,937\,529$ for Schleswig-Holstein were subdivided into the same four age strata as in the main text (see Table 1) based on census data [3]. The contact matrix in each province was estimated from Germany's contact matrix given in [4], which was aggregated to match the province's demography and symmetrized. This was done for the four contact matrices reflecting contacts at home, at work, in school, and at other locations. The matrices for home and other contacts were further split to separate the contacts between the oldest age stratum and the rest of the populations. To model contact behavior between the provinces, we hypothetically assumed that 1% of the contacts at work and at other locations are between both provinces. The exact values of the contact matrices are found in the Julia code available at GitHub (`https://github.com/Maths-against-Malaria/COVID19_Spatial_Model.git`).

From April 25, 2021 ($t = 425$) to July 4, 2021 ($t = 495$) contact reductions were incidence-based reflecting the COVID-19 emergency brake in Germany [5]. Although the same restrictions applied to both provinces, compliance with contact reducing behavior is lower in Saxony [6]. This was reflected by choosing the incidence-based reductions higher in Schleswig-Holstein (Table 2).

In both provinces, infections occurred later than in other provinces in Germany, as

**Table 1.** Age-stratified population size of Saxony (Sax) and Schleswig-Holstein (SH) chosen in simulations.

| Parameter | Description | Values | |
|---|---|---|---|
| | | Sax | SH |
| | For $a = 1, \ldots, 4$, $l = 1, 2$: | | |
| $N_{a,l}$ | Total no. of inds. in all age groups | 4 085 344 | 2 937 529 |
| $N_{1,l}$ | No. of inds. in age gr. 1 | 180 370 | 129 579 |
| $N_{2,l}$ | No. of inds. in age gr. 2 | 533 194 | 400 794 |
| $N_{3,l}$ | No. of inds. in age gr. 3 | 1 970 270 | 1 498 110 |
| $N_{4,l}$ | No. of inds. in age gr. 4 | 1 401 510 | 909 046 |

Parameters describing the population sizes. Abbreviations: inds. ... individuals.

**Table 2.** Parameters describing incidence-based contact reductions for emergency brake conditions.

| Loc. | Incidence threshold* | Home | Home(Old) | Others | Others(Old) | School | Work |
|---|---|---|---|---|---|---|---|
| Sax | <12 | 0% | 0% | 50% | 10% | 0% | 10% |
| | 12 | 25% | 5% | 60% | 20% | 50% | 55% |
| | 55 | 30% | 10% | 75% | 60% | 75% | 60% |
| | 120 | 40% | 20% | 80% | 60% | 75% | 70% |
| | 180 | 40% | 35% | 90% | 70% | 100% | 85% |
| SH | <12 | 0% | 0% | 50% | 10% | 25% | 30% |
| | 12 | 25% | 5% | 70% | 20% | 50% | 60% |
| | 55 | 30% | 10% | 85% | 60% | 75% | 65% |
| | 120 | 40% | 20% | 95% | 75% | 75% | 85% |
| | 180 | 40% | 35% | 95% | 75% | 100% | 85% |

*Incidence values (per 100 000) triggering contact reductions.

the initial cases in Germany occurred in Bavaria in late January 2020 [7] and presumably many cases were imported from travel returners from Italy into the Southern provinces in February [8] (which is the onset of the simulations presented here). Viral variants were introduced by external infections as described in the main text. Furthermore, the basic reproduction numbers and their seasonal fluctuations were chosen as described in the main text. The same applies to all parameters describing the course and severity of infections as well as the infectiveness across the course of the disease.

Vaccination campaigns started at the same times in both provinces, but the proportions of vaccinable individuals per age stratum and vaccination rates were chosen differently to reflect vaccination hesitancy in Saxony (see Tables 3 and 4).

#### Dynamics

Because the population sizes of Saxony and Schleswig-Holstein are different, we report the numbers of active infections and deaths per 100 000.

The epidemic dynamics in Saxony and Schleswig-Holstein qualitatively matched the observed dynamics in the two provinces. The first wave in both provinces was relatively mild compared to overall Germany (cf. S2 Figure).

The first wave was similar and less severe in both provinces than on average in Germany (S2 Figure A). Saxony in particular had low number of infections until the second wave. The second wave was much stronger in Saxony, exceeding the German average, while that in Schleswig-Holstein was rather low, which also lead to much lower mortality (S2 Figure B). The strong contact reductions after Christmas 2020, lead to a much higher reduction in the number of cases (in absolute and relative terms) in Saxony than in Schleswig-Holstein. The appearance of the Alpha variant caused a third wave. The fourth wave started earlier in Schleswig-Holstein than in Saxony, but so far was more successfully contained (S2 Figure A).

For both provinces a continuation of the current contact reductions would lead to strong epidemic waves. The fourth wave in Saxony would be almost 50% stronger and peak earlier than in Schleswig-Holstein (S2 Figure C,D). This is due to the contacts between the provinces. This leads to an unrealistic situation, where the model would no longer be applicable. In reality, the number of external infections would increase substantially in Schleswig-Holstein, which would lead to an earlier and stronger epidemic wave than predicted. A return to the incidence-based emergency brake at time $t = 636$ (November 22, 2021) would lead to a fast decrease in the number of cases and mortality (S2 Figure E,F).

**Table 3.** Parameters describing vaccination rates and external infections for Saxony and Schleswig-Holstein.

| Parameter | Description | | Values | |
|---|---|---|---|---|
| | Rate at which inds. get vaccinated with: | | Sax | SH |
| $\nu_{1,l}^{(1)}$ | vaccine 1 in age gr. 1 | | 0 | 0 |
| $\nu_{2,l}^{(1)}$ | vaccine 1 in age gr. 2 | $t \in [0, 590)$ | 0 | 0 |
| | | $t \in [590, 850]$ | 1/170 | 1/170 |
| $\nu_{3,l}^{(1)}$ | vaccine 1 in age gr. 3 | $t \in [0, 430)$ | 0 | 0 |
| | | $t \in [430, 590)$ | 1/240 | 1/240 |
| | | $t \in [590, 850]$ | 1/220 | 1/220 |
| $\nu_{4,l}^{(1)}$ | vaccine 1 in age gr. 4 | $t \in [0, 360)$ | 0 | 0 |
| | | $t \in [360, 400)$ | 1/280 | 1/280 |
| | | $t \in [400, 490)$ | 1/160 | 1/160 |
| | | $t \in [490, 850]$ | 1/140 | 1/140 |
| $\nu_{1,l}^{(2)}$ | vaccine 2 in age gr. 1 | | 0 | 0 |
| $\nu_{2,l}^{(2)}$ | vaccine 2 in age gr. 2 | | 0 | 0 |
| $\nu_{3,l}^{(2)}$ | vaccine 2 in age gr. 3 | $t \in [0, 430)$ | 0 | 0 |
| | | $t \in [430, 590)$ | 1/240 | 1/240 |
| | | $t \in [590, 850]$ | 1/360 | 1/360 |
| $\nu_{4,l}^{(2)}$ | vaccine 2 in age gr. 4 | $t \in [0, 400)$ | 0 | 0 |
| | | $t \in [400, 490)$ | 1/400 | 1/400 |
| | | $t \in [490, 590)$ | 1/340 | 1/340 |
| | | $t \in [590, 850]$ | 1/380 | 1/380 |
| $\nu_{1,l}^{(3)}$ | vaccine 3 in age gr. 1 | | 0 | 0 |
| $\nu_{2,l}^{(3)}$ | vaccine 3 in age gr. 2 | | 0 | 0 |
| $\nu_{3,l}^{(3)}$ | vaccine 3 in age gr. 3 | $t \in [0, 490)$ | 0 | 0 |
| | | $t \in [490, 850]$ | 1/240 | 1/240 |
| $\nu_{4,l}^{(3)}$ | vaccine 3 in age gr. 4 | $t \in [0, 490)$ | 0 | 0 |
| | | $t \in [490, 850]$ | 1/400 | 1/400 |
| $\lambda_{\text{Ext}}^{(l)}$ | Infections from outside of the population | | 1.3/day | 1/day |

Parameters describing disease progression and vaccination rates. Abbreviations: gr. . . . group.

**Table 4.** Variables describing initial values of individuals in non-infected compartments.

| Name | Description | Initial values | |
|---|---|---|---|
| | Vaccinable suscept. in: | Sax | SH |
| $S_{1,l}^{(\mathrm{U})}(0)$ | age gr. 1 | 0 | 0 |
| $S_{2,l}^{(\mathrm{U})}(0)$ | age gr. 2 | 133 299 | 140 278 |
| $S_{3,l}^{(\mathrm{U})}(0)$ | age gr. 3 | 1 379 189 | 1 348 299 |
| $S_{4,l}^{(\mathrm{U})}(0)$ | age gr. 4 | 1 191 284 | 863 594 |
| | For $a = 1, \dots, 4$, $v = 1, 2, 3, l = 1, 2$, $m = 1, 2, 3$: | | |
| $S_{a,l}^{(\mathrm{V},v)}(0)$ | vacc. suscept. with pending vaccine outcome | | 0 |
| $S_{a,l}^{(\mathrm{NI})}(0)$ | unvaccinable inds. and vaccinated suscept. that failed to immunize | | 0 |
| $S_{a,l}^{(\mathrm{PI,v})}(0)$ | vacc. suscept. who developed partial immunity | | 0 |
| $R_{a,l}^{(\mathrm{Im},v)}(0)$ | vacc. suscept. with full immunity (against at least one variant) | | 0 |
| $R_{a,l}^{(\mathrm{Inf},m)}(0)$ | recovered or fully immunized individuals | | 0 |
| $D_{a,l}^{(m)}(0)$ | individuals who die from COVID-19 | | 0 |

Summary of non-infected compartments and their initial values. Abbreviations: inds. . . . individuals; gr. . . . group; suscept. . . . susceptibles.

**Table 5.** Variables describing initial values of individuals in infected compartments.

| Name | Description ($a = 1, \ldots, 4$, $l = 1, 2$, $m = 1, 2, 3$, $v = 1, 2, 3$) | Initial Values | |
|---|---|---|---|
| | For $k = 1, \ldots, n_E$: | | |
| $E_{k,a,l}^{(\mathrm{U},m)}(0)$ | vaccinable lat. infected inds. | | 0 |
| $E_{k,a,l}^{(\mathrm{V},m,v)}(0)$ | vaccinated lat. infected inds. with pending vaccine outcome | | 0 |
| $E_{k,a,l}^{(\mathrm{NI},m)}(0)$ | unvac. & unsuccessfully immunized | | 0 |
| $E_{k,a,l}^{(\mathrm{PI},m,v)}(0)$ | vaccinated lat. infected inds. who developed partial immunity | | 0 |
| | For $k = 1, \ldots, n_P$: | | |
| $P_{k,a,l}^{(\mathrm{U},m)}(0)$ | vaccinable prod. inds. | | 0 |
| $P_{k,a,l}^{(\mathrm{V},m,v)}(0)$ | vaccinated prod. inds. with pending vaccine outcome | | 0 |
| $P_{k,a,l}^{(\mathrm{NI},m)}(0)$ | unvac. & unsuccessfully immunized prod. inds. | | 0 |
| $P_{k,a,l}^{(\mathrm{PI},m,v)}(0)$ | vaccinated prod. inds. who developed partial immunity | | 0 |
| | No. of init. infs. of vaccinable inds. in: | Sax | SH |
| $I_{1,1,l}^{(\mathrm{U},m)}(0)$ | age gr. 1 | 0 | 0 |
| $I_{1,2,l}^{(\mathrm{U},m)}(0)$ | age gr. 2 | 0 | 0 |
| $I_{1,3,l}^{(\mathrm{U},m)}(0)$ | age gr. 3 | 0.5 | 0.5 |
| $I_{1,4,l}^{(\mathrm{U},m)}(0)$ | age gr. 4 | 0 | 0 |
| | For $k = 2, \ldots, n_I$: | | |
| $I_{k,1,l}^{(\mathrm{U},m)}(0)$ | vaccinable fully inf. inds. in age gr. 1 | | 0 |
| $I_{k,2,l}^{(\mathrm{U},m)}(0)$ | vaccinable fully inf. inds. in age gr. 2 | | 0 |
| $I_{k,3,l}^{(\mathrm{U},m)}(0)$ | vaccinable fully inf. inds. in age gr. 3 | | 0 |
| $I_{k,4,l}^{(\mathrm{U},m)}(0)$ | vaccinable fully inf. inds. in age gr. 4 | | 0 |
| $I_{k,a,l}^{(\mathrm{V},m,v)}(0)$ | fully inf. inds. with pending vaccine outcome | | 0 |
| $I_{k,a,l}^{(\mathrm{NI},m)}(0)$ | unvac. & unsuccessfully immunized fully inf. inds. | | 0 |
| $I_{k,a,l}^{(\mathrm{PI},m,v)}(0)$ | Vaccinated fully inf. inds. who developed partial immunity | | 0 |
| | For $k = 2, \ldots, n_L$: | | |
| $L_{k,a,l}^{(\mathrm{U},m)}(0)$ | vaccinable late inf. individuals | | 0 |
| $L_{k,a,l}^{(\mathrm{V},m,v)}(0)$ | late inf. inds. with pending vaccine outcome | | 0 |
| $L_{k,a,l}^{(\mathrm{NI},m)}(0)$ | unvac. & unsuccessfully immunized late inf. inds. | | 0 |
| $L_{k,a,l}^{(\mathrm{PI},m,v)}(0)$ | vaccinated late inf. inds. who developed partial immunity | | 0 |

Summary of all model compartments and their initial values chosen for simulation. Abbreviations: inf. ...infectious; infs. ...infections; inds. ...individuals; lat. ...latently; prod. ...prodromal; init. ...initial; gr. ...group; unvac. ...Unvaccinable.

**Table 6.** Contact reduction parameters chosen for the simulations of Saxony.

| Parameter | Description | Home | Home(Old) | Others | Others(Old) | School | Work |
|---|---|---|---|---|---|---|---|
| gen. cont. red. | Time intervals of gen. cont. red. $t_{\text{Dist}_n}$-$t_{\text{Dist}_{n+1}}$ | | | | | | |
| $p_{\text{Cont}_1}$ | 36-85 | 25% | 5% | 80% | 20% | 90% | 75% |
| $p_{\text{Cont}_2}$ | 85-97 | 15% | 5% | 60% | 20% | 75% | 50% |
| $p_{\text{Cont}_3}$ | 97-137 | 15% | 5% | 60% | 20% | 75% | 50% |
| $p_{\text{Cont}_4}$ | 137-170 | 0% | 0% | 40% | 10% | 100% | 20% |
| $p_{\text{Cont}_5}$ | 170-180 | 0% | 0% | 40% | 10% | 100% | 20% |
| $p_{\text{Cont}_6}$ | 180-190 | 10% | 0% | 40% | 10% | 75% | 35% |
| $p_{\text{Cont}_7}$ | 190-245 | 10% | 15% | 50% | 20% | 75% | 40% |
| $p_{\text{Cont}_8}$ | 245-280 | 25% | 20% | 70% | 60% | 100% | 70% |
| $p_{\text{Cont}_9}$ | 280-303 | 30% | 20% | 80% | 60% | 100% | 75% |
| $p_{\text{Cont}_{10}}$ | 303-355 | 40% | 35% | 90% | 75% | 100% | 85% |
| $p_{\text{Cont}_{11}}$ | 355-425 | 15% | 20% | 60% | 30% | 75% | 55% |
| $p_{\text{Cont}_{12}}$ | 425-490 | 0% | 0% | 0% | 0% | 0% | 0% |
| $p_{\text{Cont}_{13}}$ | 490-540 | 10% | 5% | 50% | 30% | 50% | 25% |
| $p_{\text{Cont}_{14}}$ | 540-552 | 10% | 5% | 50% | 30% | 50% | 25% |
| $p_{\text{Cont}_{15}}$ | 552-621 | 30% | 15% | 75% | 40% | 75% | 65% |
| $p_{\text{Cont}_{16}}$ | 621-850 | 25% | 5% | 65% | 10% | 75% | 70% |

**Table 7.** Contact reduction parameters chosen for the simulations of Schleswig-Holstein.

| Parameter | Description | Home | Home(Old) | Others | Others(Old) | School | Work |
|---|---|---|---|---|---|---|---|
| gen. cont. red. | Time intervals of gen. cont. red. $t_{\mathrm{Dist}_n}$-$t_{\mathrm{Dist}_{n+1}}$ | | | | | | |
| $p_{\mathrm{Cont}_1}$ | 36-85 | 35% | 5% | 80% | 20% | 90% | 60% |
| $p_{\mathrm{Cont}_2}$ | 85-97 | 15% | 5% | 60% | 20% | 75% | 40% |
| $p_{\mathrm{Cont}_3}$ | 97-137 | 0% | 0% | 50% | 10% | 100% | 20% |
| $p_{\mathrm{Cont}_4}$ | 137-170 | 0% | 0% | 50% | 10% | 100% | 20% |
| $p_{\mathrm{Cont}_5}$ | 170-180 | 10% | 0% | 60% | 20% | 50% | 40% |
| $p_{\mathrm{Cont}_6}$ | 180-190 | 10% | 0% | 60% | 20% | 50% | 40% |
| $p_{\mathrm{Cont}_7}$ | 190-245 | 15% | 15% | 70% | 20% | 75% | 60% |
| $p_{\mathrm{Cont}_8}$ | 245-280 | 30% | 20% | 70% | 60% | 100% | 70% |
| $p_{\mathrm{Cont}_9}$ | 280-303 | 30% | 20% | 80% | 60% | 100% | 70% |
| $p_{\mathrm{Cont}_{10}}$ | 303-355 | 35% | 25% | 85% | 75% | 100% | 85% |
| $p_{\mathrm{Cont}_{11}}$ | 355-425 | 25% | 20% | 70% | 60% | 75% | 75% |
| $p_{\mathrm{Cont}_{12}}$ | 425-490 | 0% | 0% | 0% | 0% | 0% | 0% |
| $p_{\mathrm{Cont}_{13}}$ | 490-540 | 10% | 5% | 50% | 30% | 50% | 25% |
| $p_{\mathrm{Cont}_{14}}$ | 540-552 | 35% | 15% | 85% | 40% | 75% | 80% |
| $p_{\mathrm{Cont}_{15}}$ | 552-621 | 30% | 15% | 85% | 40% | 75% | 80% |
| $p_{\mathrm{Cont}_{16}}$ | 621-850 | 25% | 5% | 65% | 10% | 65% | 60% |
