## Supplementary material for "Predicting the impact of COVID-19 vaccination campaigns – a flexible age-dependent, spatially-stratified predictive model, accounting for multiple viral variants and vaccines": S1 Table

**S1 Table.** Age-stratified population size of Germany (GER).

| Parameter | Description | GER |
|---|---|---|
| $N$ | Total population size | 83 787 388 |
| $N_1$ | No. of inds. in age group 1 (0-5 years) | 3 969 138 |
| $N_2$ | No. of inds. in age group 2 (6-19 years) | 11 365 436 |
| $N_3$ | No. of inds. in age group 3 (20-59 years) | 43 730 684 |
| $N_4$ | No. of inds. in age group 4 (60+ years) | 24 722 130 |

Parameters describing the population sizes. Abbreviations: inds. . . . individuals.
