## Supplementary material for "Predicting the impact of COVID-19 vaccination campaigns – a flexible age-dependent, spatially-stratified predictive model, accounting for multiple viral variants and vaccines": S2 Table

**S2 Table.** Timeline of contact reduction measures chosen for the simulations of Germany.

| Parameter | Description | Value |
|---|---|---|
| $t_{\mathrm{Dist}_1}$ | First "hard lockdown" (general distancing) starts | 40 |
| $t_{\mathrm{Dist}_2}$ | First "hard lockdown" ends and first "relief period" starts | 85 |
| $t_{\mathrm{Dist}_3}$ | "Relief period" continues as summer vacation starts | 97 |
| $t_{\mathrm{Dist}_4}$ | "Relief period" ends as summer vacation ends | 170 |
| $t_{\mathrm{Dist}_5}$ | Start of "weak measures" after summer vacation | 190 |
| $t_{\mathrm{Dist}_6}$ | "Weak measures" ends and "soft lockdown" starts | 245 |
| $t_{\mathrm{Dist}_7}$ | "Soft lockdown" ends and Christmas "hard lockdown" starts | 280 |
| $t_{\mathrm{Dist}_8}$ | "Hard lockdown" gets stricter before Christmas | 303 |
| $t_{\mathrm{Dist}_9}$ | "Hard lockdown" ends and "soft lockdown" starts | 355 |
| $t_{\mathrm{Dist}_{10}}$ | Start of "emergency brake" on April 25 | 425 |
| $t_{\mathrm{Dist}_{11}}$ | "emergency brake" ends | 490 |
| $t_{\mathrm{Dist}_{12}}$ | "Soft lockdown" and 3G rule starts | 540 |
| $t_{\mathrm{Dist}_{13}}$ | "Soft lockdown" ends and"relief period" and 2G rule starts | 621 |
| $t_{\mathrm{Dist}_{14}}$ | "Hypothetical school closures" starts | 636 |
| $t_{\mathrm{Dist}_{15}}$ | "Hypothetical school closures" ends | 850 |
