## Supplementary material for "Predicting the impact of COVID-19 vaccination campaigns – a flexible age-dependent, spatially-stratified predictive model, accounting for multiple viral variants and vaccines": S3 Table

**S3 Table.** Contact reduction parameters chosen for the simulations.

| Parameter | Description | Home | Home(Old) | Others | Others(Old) | School | Work |
|---|---|---|---|---|---|---|---|
| gen. cont. red. | Time intervals of gen. cont. red. $t_{\text{Dist}_n}$-$t_{\text{Dist}_{n+1}}$ | | | | | | |
| $p_{\text{Cont}_1}$ | 40-85 | 35% | 5% | 95% | 20% | 95% | 70% |
| $p_{\text{Cont}_2}$ | 85-97 | 15% | 5% | 60% | 20% | 75% | 50% |
| $p_{\text{Cont}_3}$ | 97-170 | 0% | 0% | 40% | 10% | 100% | 20% |
| $p_{\text{Cont}_4}$ | 170-190 | 10% | 0% | 50% | 10% | 50% | 40% |
| $p_{\text{Cont}_5}$ | 190-245 | 15% | 5% | 70% | 20% | 75% | 50% |
| $p_{\text{Cont}_6}$ | 245-280 | 33% | 30% | 80% | 70% | 100% | 75% |
| $p_{\text{Cont}_7}$ | 280-303 | 35% | 30% | 80% | 75% | 100% | 75% |
| $p_{\text{Cont}_8}$ | 303-355 | 30% | 35% | 95% | 75% | 100% | 85% |
| $p_{\text{Cont}_9}$ | 355-425 | 25% | 20% | 75% | 60% | 75% | 60% |
| $p_{\text{Cont}_{10}}$ | 425-490 | 0% | 0% | 0% | 0% | 0% | 0% |
| $p_{\text{Cont}_{11}}$ | 490-540 | 10% | 5% | 40% | 25% | 50% | 20% |
| $p_{\text{Cont}_{12}}$ | 540-621 | 30% | 15% | 85% | 40% | 75% | 65% |
| $p_{\text{Cont}_{13}}$ | 621-636 | 30% | 15% | 85% | 40% | 75% | 80% |
| $p_{\text{Cont}_{14}}$ | 636-850 | 30% | 15% | 85% | 40% | 75% | 80% |
