## Supplementary material for "Predicting the impact of COVID-19 vaccination campaigns – a flexible age-dependent, spatially-stratified predictive model, accounting for multiple viral variants and vaccines": S4 Table

**S4 Table.** Parameters describing incidence-based contact reductions for emergency brake conditions.

| Incidence thresholds* | Home | Home(Old) | Others | Others(Old) | School | Work |
|---|---|---|---|---|---|---|
| <10 | 0% | 0% | 50% | 10% | 25% | 30% |
| 10 | 25% | 5% | 60% | 20% | 50% | 55% |
| 50 | 30% | 10% | 85% | 60% | 75% | 65% |
| 100 | 40% | 20% | 95% | 75% | 75% | 85% |
| 180 | 40% | 35% | 95% | 75% | 100% | 85% |

*Incidence values (per 100 000) triggering contact reductions.
