## Supplementary material for "Predicting the impact of COVID-19 vaccination campaigns – a flexible age-dependent, spatially-stratified predictive model, accounting for multiple viral variants and vaccines": S5 Table

**S5 Table.** Parameters describing the vaccination rate.

| Parameter | Description | | Value |
|---|---|---|---|
| | Rate at which inds. get vaccinated with: | | |
| $\nu_{1,l}^{(1)}$ | vaccine 1 in age gr. 1 | | 0 |
| $\nu_{2,l}^{(1)}$ | vaccine 1 in age gr. 2 | $t \in [0, 590)$ | 0 |
| | | $t \in [590, 850]$ | 1/170 |
| $\nu_{3,l}^{(1)}$ | vaccine 1 in age gr. 3 | $t \in [0, 430)$ | 0 |
| | | $t \in [430, 850]$ | 1/240 |
| $\nu_{4,l}^{(1)}$ | vaccine 1 in age gr. 4 | $t \in [0, 360)$ | 0 |
| | | $t \in [360, 400)$ | 1/280 |
| | | $t \in [400, 490)$ | 1/160 |
| | | $t \in [490, 850]$ | 1/140 |
| $\nu_{1,l}^{(2)}$ | vaccine 2 in age gr. 1 | | 0 |
| $\nu_{2,l}^{(2)}$ | vaccine 2 in age gr. 2 | | 0 |
| $\nu_{3,l}^{(2)}$ | vaccine 2 in age gr. 3 | $t \in [0, 430)$ | 0 |
| | | $t \in [430, 850]$ | 1/240 |
| $\nu_{4,l}^{(2)}$ | vaccine 2 in age gr. 4 | $t \in [0, 400)$ | 0 |
| | | $t \in [400, 490)$ | 1/400 |
| | | $t \in [490, 850]$ | 1/340 |
| $\nu_{1,l}^{(3)}$ | vaccine 3 in age gr. 1 | | 0 |
| $\nu_{2,l}^{(3)}$ | vaccine 3 in age gr. 2 | | 0 |
| $\nu_{3,l}^{(3)}$ | vaccine 3 in age gr. 3 | $t \in [0, 490)$ | 0 |
| | | $t \in [490, 850]$ | 1/240 |
| $\nu_{4,l}^{(3)}$ | vaccine 3 in age gr. 4 | $t \in [0, 490)$ | 0 |
| | | $t \in [490, 850]$ | 1/400 |

Parameters describing vaccination rates and their values used in the simulations. Abbreviations: gr. . . . group.
