## Supplementary material for "Predicting the impact of COVID-19 vaccination campaigns – a flexible age-dependent, spatially-stratified predictive model, accounting for multiple viral variants and vaccines": S6 Table

**S6 Table.** Parameters describing disease progression.

| Parameter | Description | Value |
|---|---|---|
| $n_E$ | No. of latent sub-states (Erlang states) | 5 |
| $n_P$ | No. of prodromal sub-states (Erlang states) | 5 |
| $n_I$ | No. of fully-infectious sub-states (Erlang states) | 5 |
| $n_L$ | No. of late-infectious sub-states (Erlang states) | 5 |
| | Average duration of: | |
| $D_E$ | latent period | 3.5 days |
| $D_P$ | prodromal period | 1 day |
| $D_I$ | fully-infectious period | 5 days |
| $D_L$ | late-infectious period | 5 days |
| | Transition rate of ($a = 1, \ldots, 4$, $m = 1, 2, 3$): | |
| $\varepsilon_{a,m}$ | latent sub-states | $n_{\mathrm{E}}/D_{\mathrm{E}}$ |
| $\varphi_{a,m}$ | prodromal sub-states | $n_{\mathrm{P}}/D_{\mathrm{P}}$ |
| $\gamma_{a,m}$ | fully-infectious sub-states | $n_{\mathrm{I}}/D_{\mathrm{I}}$ |
| $\delta_{a,m}$ | late-infectious sub-states | $n_{\mathrm{L}}/D_{\mathrm{L}}$ |
| | For $a = 1, \ldots, 4$: | |
| $\alpha_a^{(1)}, \alpha_a^{(2)}, \alpha_a^{(3)}$ | average waiting time for the outcome of the vaccines | $1/30, 1/50, 1/15$ |
