## Supplementary material for "Predicting the impact of COVID-19 vaccination campaigns – a flexible age-dependent, spatially-stratified predictive model, accounting for multiple viral variants and vaccines": S7 Table

**S7 Table.** Parameters describing vaccination outcome and immunity.

| Parameters | Description ($m = 1, 2, 3$) | Value | | |
|---|---|---|---|---|
| | Fraction of susceptibles that: $v$: | 1 | 2 | 3 |
| $f_S^{(\mathrm{NI},v)}$ | fail to immunize upon vaccination | 0.05 | 0.05 | 0.05 |
| $f_S^{(\mathrm{PI},v)}$ | develop partial immunity upon vaccination | 0.70 | 0.65 | 0.65 |
| $f_S^{(\mathrm{Im},v)}$ | develop full immunity after vaccination | 0.25 | 0.30 | 0.30 |
| | Fraction of latent individuals that: | | | |
| $f_E^{(\mathrm{NI},v)}$ | fail to immunize after vaccination | 0.50 | 0.50 | 0.50 |
| $f_E^{(\mathrm{PI},v)}$ | develop partial immunity after vaccination | 0.50 | 0.50 | 0.50 |
| $f_E^{(\mathrm{Im},v)}$ | develop full immunity after vaccination | 0.00 | 0.00 | 0.00 |
| | Fraction of prodromal individuals that: | | | |
| $f_P^{(\mathrm{NI},v)}$ | fail to immunize after vaccination | 0.50 | 0.50 | 0.50 |
| $f_P^{(\mathrm{PI},v)}$ | develop partial immunity after vaccination | 0.50 | 0.50 | 0.50 |
| $f_P^{(\mathrm{Im},v)}$ | develop full immunity after vaccination | 0.00 | 0.00 | 0.00 |
| | Fraction of fully-infectious individuals that: | | | |
| $f_I^{(\mathrm{NI},v)}$ | fail to immunize after vaccination | 0.50 | 0.50 | 0.50 |
| $f_I^{(\mathrm{PI},v)}$ | develop partial immunity after vaccination | 0.50 | 0.50 | 0.50 |
| $f_I^{(\mathrm{Im},v)}$ | develop full immunity after vaccination | 0.00 | 0.00 | 0.00 |
| | Fraction of late-infectious individuals that: | | | |
| $f_L^{(\mathrm{NI},v)}$ | fail to immunize after vaccination | 0.50 | 0.50 | 0.50 |
| $f_L^{(\mathrm{PI},v)}$ | develop partial immunity after vaccination | 0.50 | 0.50 | 0.50 |
| $f_L^{(\mathrm{Im},v)}$ | develop full immunity after vaccination | 0.00 | 0.00 | 0.00 |
| | Fraction by which partial immunity reduces transmissibility in the: | | | |
| $p_P^{(m,v)}$ | prodromal period | 0.66 | 0.70 | 0.66 |
| $p_I^{(m,v)}$ | fully-infectious period | 0.66 | 0.70 | 0.66 |
| $p_L^{(m,v)}$ | late-infectious period | 0.66 | 0.70 | 0.66 |
| | Susceptibility reduced by partial immunity against: | | | |
| $g(1, v)$ | variant 1 | 0.50 | 0.50 | 0.55 |
| $g(2, v)$ | variant 2 | 0.60 | 0.60 | 0.60 |
| $g(3, v)$ | variant 3 | 0.60 | 0.60 | 0.60 |
| $h(m, v)$ | Susceptibility reduced by full immunity | | | 0 |
