## Supplementary material for "Predicting the impact of COVID-19 vaccination campaigns – a flexible age-dependent, spatially-stratified predictive model, accounting for multiple viral variants and vaccines": S8 Table

**S8 Table.** Parameters describing disease severity and mortality.

| Parameters | Description ($m = 1, 2, 3$) | | Value | | |
|---|---|---|---|---|---|
| | Fraction of sympt. inds. in: | $m$: | 1 | 2 | 3 |
| $f_{\text{Sick}}^{(1,m)}$ | age gr. 1 | | 0.15 | 0.15 | 0.15 |
| $f_{\text{Sick}}^{(2,m)}$ | age gr. 2 | | 0.30 | 0.30 | 0.30 |
| $f_{\text{Sick}}^{(3,m)}$ | age gr. 3 | | 0.65 | 0.65 | 0.65 |
| $f_{\text{Sick}}^{(4,m)}$ | age gr. 4 | | 0.70 | 0.70 | 0.70 |
| | Fraction of sympt. inds. who die in: | $m$: | 1 | 2 | 3 |
| $f_{\text{dead}}^{(1,m)}$ | age gr. 1 | | 0.0001 | 0.0002 | 0.0004 |
| $f_{\text{dead}}^{(2,m)}$ | age gr. 2 | | 0.0001 | 0.0002 | 0.0004 |
| $f_{\text{dead}}^{(3,m)}$ | age gr. 3 | | 0.0010 | 0.0011 | 0.0012 |
| $f_{\text{dead}}^{(4,m)}$ | age gr. 4 | | 0.2400 | 0.2400 | 0.2400 |
| | Fraction of sympt. inds. with part. immunity in: | $v$: | 1 | 2 | 3 |
| $f_{\text{Sick}}^{(\text{PI},1,1,v)}$ | age gr. 1 infected with variant 1 | | 0.10 | 0.11 | 0.10 |
| $f_{\text{Sick}}^{(\text{PI},1,2,v)}$ | age gr. 1, infected with variant 2 | | 0.11 | 0.12 | 0.11 |
| $f_{\text{Sick}}^{(\text{PI},1,3,v)}$ | age gr. 1, infected with variant 3 | | 0.12 | 0.13 | 0.12 |
| $f_{\text{Sick}}^{(\text{PI},2,1,v)}$ | age gr. 2, infected with variant 1 | | 0.25 | 0.22 | 0.25 |
| $f_{\text{Sick}}^{(\text{PI},2,2,v)}$ | age gr. 2, infected with variant 2 | | 0.26 | 0.23 | 0.26 |
| $f_{\text{Sick}}^{(\text{PI},2,3,v)}$ | age gr. 2, infected with variant 3 | | 0.27 | 0.24 | 0.27 |
| $f_{\text{Sick}}^{(\text{PI},3,1,v)}$ | age gr. 3, infected with variant 1 | | 0.55 | 0.60 | 0.55 |
| $f_{\text{Sick}}^{(\text{PI},3,2,v)}$ | age gr. 3, infected with variant 2 | | 0.56 | 0.61 | 0.56 |
| $f_{\text{Sick}}^{(\text{PI},3,3,v)}$ | age gr. 3, infected with variant 3 | | 0.57 | 0.62 | 0.57 |
| $f_{\text{Sick}}^{(\text{PI},4,1,v)}$ | age gr. 4, infected with variant 1 | | 0.60 | 0.60 | 0.60 |
| $f_{\text{Sick}}^{(\text{PI},4,2,v)}$ | age gr. 4, infected with variant 2 | | 0.60 | 0.60 | 0.60 |
| $f_{\text{Sick}}^{(\text{PI},4,3,v)}$ | age gr. 4, infected with variant 3 | | 0.60 | 0.60 | 0.60 |
| | Fraction of inds. with part. immunity who die in: | $v$: | 1 | 2 | 3 |
| $f_{\text{Dead}}^{(\text{PI},1,1,v)}$ | age gr. 1, infected with variant 1 | | 0.00 | 0.00 | 0.00 |
| $f_{\text{Dead}}^{(\text{PI},1,2,v)}$ | age gr. 1, infected with variant 2 | | 0.00 | 0.00 | 0.00 |
| $f_{\text{Dead}}^{(\text{PI},1,3,v)}$ | age gr. 1, infected with variant 3 | | 0.00 | 0.00 | 0.00 |
| $f_{\text{Dead}}^{(\text{PI},2,1,v)}$ | age gr. 2, infected with variant 1 | | 0.5e-5 | 5.5e-6 | 0.5e-5 |
| $f_{\text{Dead}}^{(\text{PI},2,2,v)}$ | age gr. 2, infected with variant 2 | | 1.0e-5 | 1.6e-5 | 1.0e-5 |
| $f_{\text{Dead}}^{(\text{PI},2,3,v)}$ | age gr. 2, infected with variant 3 | | 2.0e-5 | 2.5e-5 | 2.0e-5 |
| $f_{\text{Dead}}^{(\text{PI},3,1,v)}$ | age gr. 3, infected with variant 1 | | 5.5e-5 | 6.0e-5 | 5.5e-5 |
| $f_{\text{Dead}}^{(\text{PI},3,2,v)}$ | age gr. 3, infected with variant 2 | | 5.5e-5 | 6.0e-5 | 5.5e-5 |
| $f_{\text{Dead}}^{(\text{PI},3,3,v)}$ | age gr. 3, infected with variant 3 | | 5.5e-5 | 6.0e-5 | 5.5e-5 |
| $f_{\text{Dead}}^{(\text{PI},4,1,v)}$ | age gr. 4, infected with variant 1 | | 0.0040 | 0.0045 | 0.0040 |
| $f_{\text{Dead}}^{(\text{PI},4,2,v)}$ | age gr. 4, infected with variant 2 | | 0.0040 | 0.0045 | 0.0040 |
| $f_{\text{Dead}}^{(\text{PI},4,3,v)}$ | age gr. 4, infected with variant 3 | | 0.0040 | 0.0045 | 0.0040 |

Summary of parameters describing disease severity and mortality. Abbreviations: inds. . . . individuals; sympt. . . . symptomatic.
