## Supplementary material for "Predicting the impact of COVID-19 vaccination campaigns – a flexible age-dependent, spatially-stratified predictive model, accounting for multiple viral variants and vaccines": S9 Table

**S9 Table.** Variables describing initial values of individuals in non-infected compartments.

| Name | Description | Initial Values |
|---|---|---|
| | Vaccinable suscept. in: | |
| $S_1^{(\mathrm{U})}(0)$ | age group 1 | 0 |
| $S_2^{(\mathrm{U})}(0)$ | age group 2 | 4 091 557 |
| $S_3^{(\mathrm{U})}(0)$ | age group 3 | 37 171 081 |
| $S_4^{(\mathrm{U})}(0)$ | age group 4 | 23 486 024 |
| | For $a = 1, \ldots, 4$, $v = 1, 2, 3$, $m = 1, 2, 3$: | |
| $S_a^{(\mathrm{V},v)}(0)$ | vacc. suscept. with pending vaccine outcome | 0 |
| $S_a^{(\mathrm{NI})}(0)$ | unvaccinable & unsuccessfully immune inds. | 0 |
| $S_a^{(\mathrm{PI},v)}(0)$ | vacc. suscept. who developed partial immunity | 0 |
| $R_a^{(\mathrm{Im},v)}(0)$ | vacc. suscept. with full immunity (against at least one variant) | 0 |
| $R_a^{(\mathrm{Inf},m)}(0)$ | Recovered or fully immune individuals | 0 |
| $D_a^{(m)}(0)$ | Individuals who die from COVID-19 | 0 |

Summary of non-infected compartments and their initial values. Abbreviations: inds. . . . individuals; suscept. . . . susceptibles; vacc. . . . vaccinated.
