## Supplementary material for "Predicting the impact of COVID-19 vaccination campaigns – a flexible age-dependent, spatially-stratified predictive model, accounting for multiple viral variants and vaccines": S10 Table

**S10 Table.** Variables describing initial values of individuals in infected compartments.

| Name | Description ($a = 1, \ldots, 4$, $m = 1, 2, 3$, $v = 1, 2, 3$) | Initial Values |
|---|---|---|
| | For $k = 1, \ldots, n_E$: | |
| $E_{k,a}^{(\mathrm{U},m)}(0)$ | vaccinable lat. infected inds. | 0 |
| $E_{k,a}^{(\mathrm{V},m,v)}(0)$ | vaccinated lat. infected inds. with pending vaccine outcome | 0 |
| $E_{k,a}^{(\mathrm{NI},m)}(0)$ | unvaccinable & unsuccessfully lat. infected inds. | 0 |
| $E_{k,a}^{(\mathrm{PI},m,v)}(0)$ | vaccinated lat. infected inds. who developed partial immunity | 0 |
| | For $k = 1, \ldots, n_P$: | |
| $P_{k,a}^{(\mathrm{U},m)}(0)$ | vaccinable prodromal inds. | 0 |
| $P_{k,a}^{(\mathrm{V},m,v)}(0)$ | vaccinated prodromal inds. with pending vaccine outcome | 0 |
| $P_{k,a}^{(\mathrm{NI},m)}(0)$ | unvaccinable & unsuccessfully immune prodromal inds. | 0 |
| $P_{k,a}^{(\mathrm{PI},m,v)}(0)$ | vaccinated prodromal inds. who developed partial immunity | 0 |
| | No. of initial infs. of vaccinable inds. in: | |
| $I_{1,1}^{(\mathrm{U},m)}(0)$ | age gr. 1 | 0 |
| $I_{1,2}^{(\mathrm{U},m)}(0)$ | age gr. 2 | 10 |
| $I_{1,3}^{(\mathrm{U},m)}(0)$ | age gr. 3 | 50 |
| $I_{1,4}^{(\mathrm{U},m)}(0)$ | age gr. 4 | 5 |
| | For $k = 2, \ldots, n_I$: vaccinable fully-infectious inds. in: | |
| $I_{k,1}^{(\mathrm{U},m)}(0)$ | age gr. 1 | 0 |
| $I_{k,2}^{(\mathrm{U},m)}(0)$ | age gr. 2 | 0 |
| $I_{k,3}^{(\mathrm{U},m)}(0)$ | age gr. 3 | 0 |
| $I_{k,4}^{(\mathrm{U},m)}(0)$ | age gr. 4 | 0 |
| $I_{k,a}^{(\mathrm{V},m,v)}(0)$ | fully-infectious inds. with pending vaccine outcome | 0 |
| $I_{k,a}^{(\mathrm{NI},m)}(0)$ | unvaccinable & unsuccessfully immune fully-infectious inds. | 0 |
| $I_{k,a}^{(\mathrm{PI},m,v)}(0)$ | vaccinated fully-infectious inds. who developed partial immunity | 0 |
| | For $k = 2, \ldots, n_L$: | |
| $L_{k,a}^{(\mathrm{U},m)}(0)$ | vaccinable late-infectious inds. | 0 |
| $L_{k,a}^{(\mathrm{V},m,v)}(0)$ | late-infectious inds. with pending vaccine outcome | 0 |
| $L_{k,a}^{(\mathrm{NI},m)}(0)$ | unvaccinable & unsuccessfully immune late-infectious inds. | 0 |
| $L_{k,a}^{(\mathrm{PI},m,v)}(0)$ | vaccinated late-infectious inds. who developed partial immunity | 0 |

Summary of infected compartments and their initial values chosen for simulation. Abbreviations: inds. ... individuals; lat. ... latently; gr. ... group.
