## Supplementary material for "Predicting the impact of COVID-19 vaccination campaigns – a flexible age-dependent, spatially-stratified predictive model, accounting for multiple viral variants and vaccines": S11 Table

**S11 Table.** Parameters describing external-, seasonal-factors, control measures, and contagiousness.

| Parameter | Definition | Value/Eq. |
|---|---|---|
| $\lambda_{\text{Ext}}$ | Infections from outside of the population | 30/day |
| $\bar{R}_0^{(1)}, \bar{R}_0^{(2)}, \bar{R}_0^{(3)}$ | Annual average basic reproduction number | 3.0, 3.9, 5.46 |
| $A$ | Amplitude of the seasonal fluctuation in $R_0$ | 0.43 |
| $t_{R_{0_{\max}}}$ | Day when $R_0$ reaches its maximum | 300 |
| $Q_{\max}$ | Maximum capacity of isolation units per 10,000 | 200 |
| $t_{\text{Iso}_2}$ | Day case isolation measures start | 10 |
| $t_{\text{Iso}_1}$ | Day case isolation measures end | 850 |
| $t_{\text{Mut}}^{(1)}, t_{\text{Mut}}^{(2)}, t_{\text{Mut}}^{(3)}$ | Day mutations are introduced into the population | -20, 290, 475 |
| $f_{\text{Iso}}$ | Fraction of inf. inds. who are isolated | 58% |
| $p_{\text{Home}}$ | Contact reduction in home isolation | 75% |
| $c_P$ | Relative contagiousness in prodromal period | 0.5 |
| $c_I$ | Relative contagiousness in fully-infectious phase | 1 |
| $c_L$ | Relative contagiousness in late-infectious phase | 0.5 |
| $\beta_P(t)$ | Seasonally varying effective contact rate of prodromal inds. | cf. eq. 8a |
| $\beta_I(t)$ | Seasonally varying effective contact rate of fully-infectious inds. | cf. eq. 8b |
| $\beta_L(t)$ | Seasonally varying effective contact rate of late-infectious inds. | cf. eq. 8c |

| | | | | |
|---|---|---|---|---|
| | Day when vaccination campaigns start for: | $v$: 1 | 2 | 3 |
| $t_{\text{Vacc}}^{(1,v)}$ | age group 1 | 850 | 850 | 850 |
| $t_{\text{Vacc}}^{(2,v)}$ | age group 2 | 490 | 850 | 850 |
| $t_{\text{Vacc}}^{(3,v)}$ | age group 3 | 400 | 400 | 430 |
| $t_{\text{Vacc}}^{(4,v)}$ | age group 4 | 310 | 360 | 430 |
