## Supplementary material for "Predicting the impact of COVID-19 vaccination campaigns – a flexible age-dependent, spatially-stratified predictive model, accounting for multiple viral variants and vaccines": S1 Figure

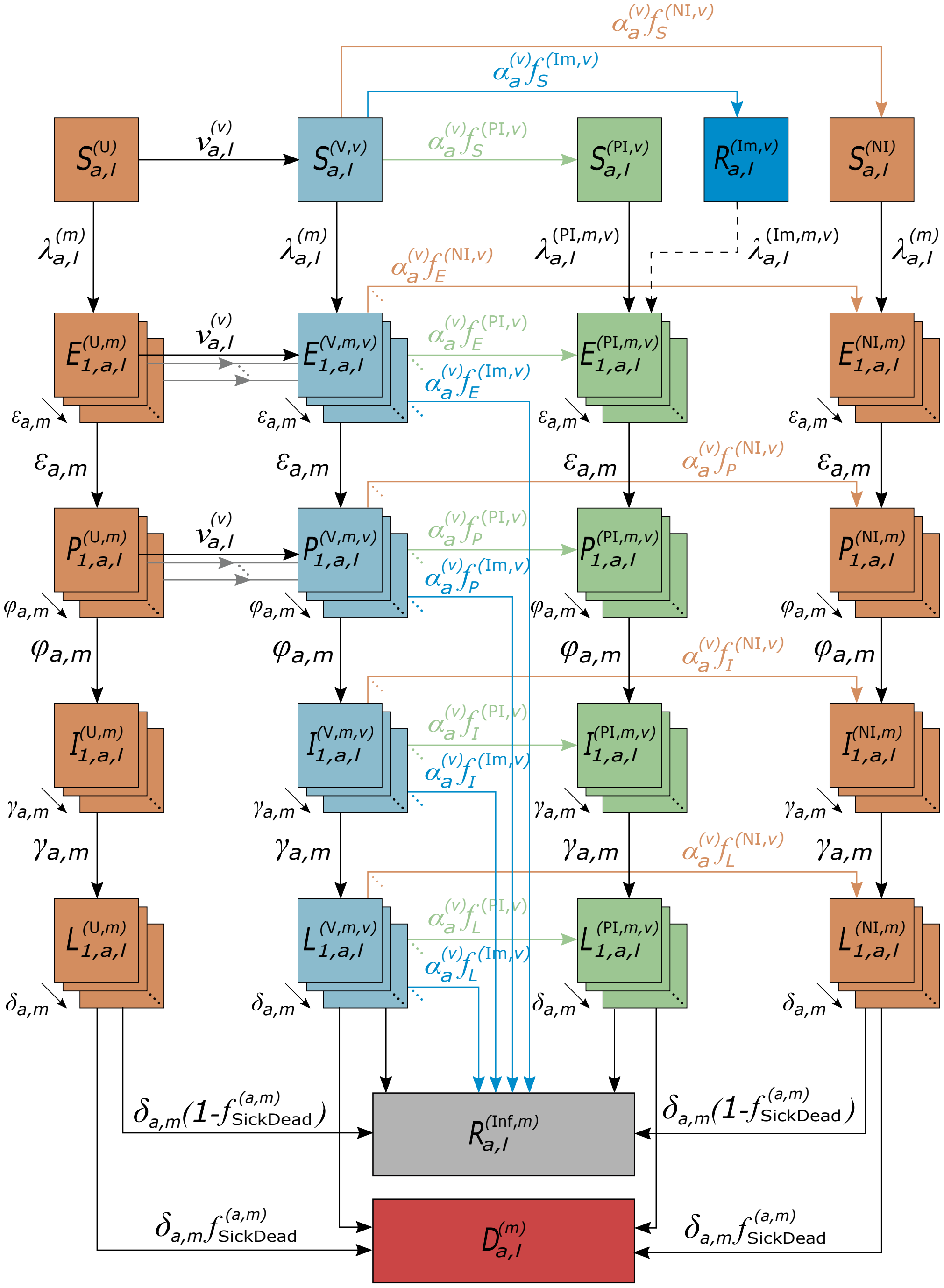


$$f_{\mathrm{SickDead}}^{(a,m)} = f_{\mathrm{Sick}}^{(a,m)} f_{\mathrm{Dead}}^{(a,m)}$$

**Fig 1. Flow chart:** in the flow chart, Greek letters indicate rates. Capital letters $S$, $E$, $P$, $I$, $L$, $R$ and $D$ stand for susceptible, latent (early), prodromal, fully-infectious, late-infectious, recovered, and dead, respectively. Overlapping squares indicate equivalent compartments, which are surpassed successively (the number 1 in the sub-script indicates the first of the respective sub-states). The sub-scripts $a$ and $l$ indicate the age group and location. In the super-scripts, $m$ indicates the infecting variant and $v$ the different vaccines. Moreover, in the super-scripts 'U' indicated unvaccinated individuals waiting to be vaccinated , 'V' indicates vaccinated individuals for which the outcome of the vaccine is pending, 'PI' partially-immune individuals, and 'NI' individuals that are unvaccinable or failed to immunize. Finally, 'Inf' indicates that individuals recovered from infection, and 'Im' implies that individuals were completely immunized against at least one variant.
