## Supplementary material for "Predicting the impact of COVID-19 vaccination campaigns – a flexible age-dependent, spatially-stratified predictive model, accounting for multiple viral variants and vaccines": S2 Figure

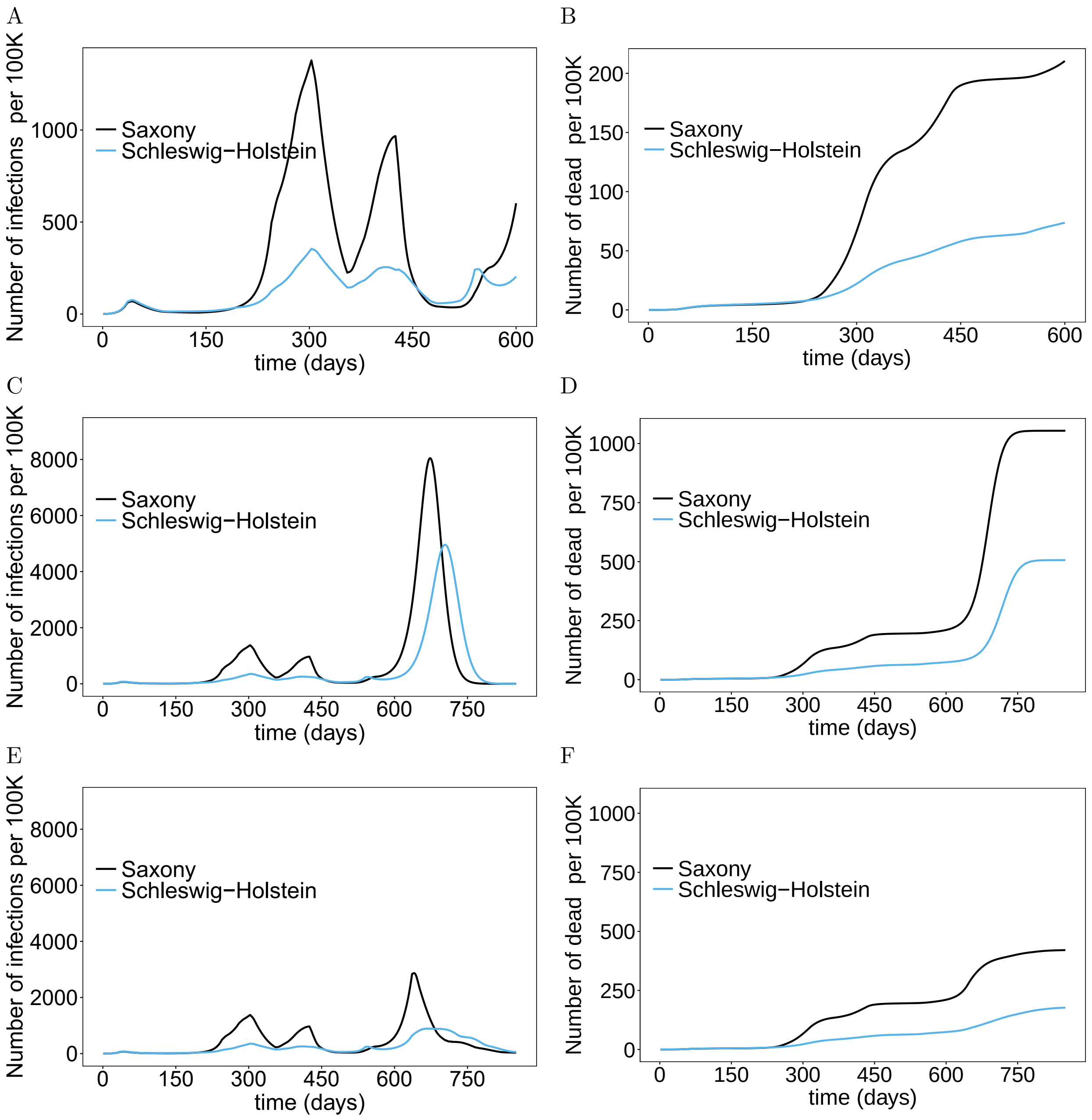


**S2 Fig. Effect of emergency brake – two locations model:** (A) Shown are the total numbers of infected individuals assuming the same contact reductions after November 7, 2021 as one year earlier, with and without the weather adjustments in 2021. (B) Shown are the corresponding deaths. The parameters used for the simulations are listed in S1 Table-S4 Table.
